# Reservoir and Phylogenetic Signatures Identify Distinct Subsets of HIV-1 Nonsuppressible Viremia

**DOI:** 10.64898/2026.02.05.26345678

**Authors:** Taylor Adams, Chang Kyung Kang, Abbas Mohammadi, Flavio Mesquita, Sofia Cohen, Efthimios A. Deligiannidis, Gregory E. Edelstein, Dominic Dorazio, Antonia de Andraca Serrano, Julian Kim, Matthew Moeser, Lindsey E. Hastings, Liam Carvalho, Hannah Jordan, Daniel P. Worrall, Jose R. Castillo-Mancilla, Nikolaus Jilg, Jeffrey M. Jacobson, Athe N. Tsibris, Steven Deeks, Courtney Fletcher, Josep M. Llibre, Peter L. Anderson, Shuntai Zhou, Sarah B. Joseph, Scott Sieg, Steven Yukl, Behzad Etemad, Jonathan Z. Li

**Affiliations:** Brigham and Women’s Hospital, Harvard Medical School, Boston, MA, USA; Seoul National University College of Medicine, Seoul, Republic of Korea; Valley Health System, Las Vegas, NV, USA; Division of Infectious Diseases and HIV Medicine, Department of Medicine, Case Western Reserve University/University Hospitals Cleveland Medical Center, Cleveland, OH, USA; Division of HIV, Infectious Diseases, and Global Medicine, University of California, San Francisco, CA, USA; Department of Microbiology and Immunology, University of North Carolina at Chapel Hill, Chapel Hill, NC, USA; Division of Pediatric Infectious Diseases, Department of Pediatrics, University of Alabama at Birmingham, Birmingham, AL, USA; Ragon Institute of MGH, MIT, and Harvard, Cambridge, MA, USA; Skaggs School of Pharmacy and Pharmaceutical Sciences, University of Colorado Anschutz Medical Campus, Aurora, CO, USA; Division of Infectious Diseases, Department of Medicine, University of Colorado Anschutz Medical Campus, Aurora, CO, USA; Antiviral Pharmacology Laboratory, College of Pharmacy, University of Nebraska Medical Center, 986145 Nebraska Medical Center, Omaha, NE, USA; Division of Infectious Diseases, Department of Medicine, University of Nebraska Medical Center, Omaha, NE, USA; Division of Infectious Diseases, Hospital Universitari Germans Trias i Pujol, Barcelona, Spain

## Abstract

In nonsuppressible HIV viremia (NSV), individuals have persistently detectable viral load despite adherence to ≥2 fully active antiretroviral drugs. NSV represents an area of clinical uncertainty and an opportunity to understand the mechanisms of HIV persistence. We performed in-depth virologic characterization to identify distinct NSV phenotypes. We categorized participants into those who had persistent viremia after antiretroviral therapy (ART) initiation (primary NSV) and those who had NSV after a period of virologic suppression (secondary NSV). Despite the prolonged viremia, there was no significant evidence of active viral evolution in either the primary or secondary NSV groups. Primary NSV participants had >10-fold higher levels of intact proviral DNA by the intact proviral DNA assay (*P*<0.01). While the plasma of secondary NSV participants was dominated by a few large HIV clones, primary NSV participants had far more diverse plasma quasispecies with few clones (*P*<0.01). Primary NSV participants were also found to harbor distinct deletions within *vif-vpr* and had T-tropic virus. Transcriptional profiling of intracellular HIV RNA also suggested higher viral transcriptional activity in primary than in secondary NSV. In contrast, profiling of soluble inflammatory markers demonstrated largely comparable systemic inflammatory signatures across NSV subtypes. NSV is comprised of two distinct subsets of individuals, including a novel group with primary NSV characterized by prolonged viremia after ART initiation, an exceptionally large intact reservoir and highly diverse plasma virus populations arising from transcriptionally active proviral reservoirs, without evidence of ongoing evolution. These findings have implications for understanding mechanisms of HIV reservoir persistence on ART.

**One Sentence Summary:** Two distinct subsets of HIV-1 nonsuppressible viremia, primary and secondary, are identified and characterized by reservoir and phylogenetic characteristics.

## Introduction

Although antiretroviral therapy (ART) has achieved durable viral suppression in most people living with human immunodeficiency virus (HIV), a subset of individuals experience persistent low-level viremia (pLLV) despite ART. While suboptimal ART adherence or drug resistance has traditionally been thought to be the cause of pLLV, a subset of these patients have been found to have non-suppressible HIV viremia (NSV) caused by a hyperactive HIV reservoir and in the absence of ART non-adherence or drug resistance (*1–3*). NSV has been increasingly recognized as a clinical challenge and an extreme example of the barriers to viral reservoir control and HIV cure efforts. As such, a detailed understanding of NSV offers a unique opportunity to probe the mechanisms of HIV persistence.

While ART effectively controls plasma viremia in most people with HIV (PWH), the latent reservoir remains a key driver of HIV persistence. This reservoir exhibits dynamic stability, which reflects a balance between the natural decay of infected cells and their replenishment through clonal expansion and long-term survival (*4–7*). Such expanded clones can periodically produce virus, resulting in low-level viremia even under fully suppressive ART, and recent evidence indicates that the same mechanism underlies NSV. Mohammadi *et al.* demonstrated that NSV is predominantly driven by clonally expanded CD4⁺ T cells that can produce infectious virus (*1*). They showed that persistent viremia during ART often originates from identical proviral sequences linked to specific integration sites, indicating reactivation of expanded clones rather than ongoing viral replication.

However, prior NSV studies have almost exclusively focused on cases where individuals first achieved viral suppression and subsequently developed persistent viremia. In contrast, little is known about individuals who never achieve viral suppression despite continuous ART. This phenotype has occasionally been described in observational studies (*8*), but a recent review has emphasized its rarity and uncertain biology (*9*). This unique clinical form of NSV, which we term primary NSV, is frequently accompanied by viral loads that can exceed 1,000 HIV copies/mL and may be characterized by features distinct from previously described secondary NSV participants who were able to maintain viral suppression prior to the NSV.

In this study, we performed a detailed assessment of reservoir size, plasma and proviral composition by single-genome near-full length sequencing, phylogenetic structure and diversity, HIV cellular tropism, HIV RNA transcriptional profile, and inflammatory environment. This work alters the current paradigm, providing new insights into the heterogeneity of NSV, and has implications for understanding HIV persistence and guiding future cure strategies.

## Results

### NSV participant characteristics

Since our description of eight individuals with NSV who had initially achieved viral suppression before developing persistent viremia (*1*), we have expanded the cohort to include 15 total participants, including 11 men (73%), with a median age at study entry of 55 years (Table S1). The median duration of ART was 13.9 years (range, 3.0–27.9). Median CD4⁺ T-cell nadir, defined as the lowest documented CD4⁺ T-cell count, and the most recent CD4⁺ T-cell count were 208 cells/mm³ and 647 cells/mm³, respectively. ART regimens and the genotypic susceptibility scores (GSS) of plasma viruses sequenced at study entry are summarized in Table S1. All participants had been receiving at least two fully active antiretroviral drugs, which was consistent with viremia due to NSV, rather than replication of drug-resistant virus.

Interestingly, three of the newly enrolled participants with NSV were unable to achieve viral suppression at any time point after initiating ART. This clinical presentation was clearly distinct from the other NSV participants, all of whom had maintained periods of virologic suppression after ART initiation. Based on this observation, we classified cases into two groups: primary NSV, defined as NSV in individuals who had never been suppressed despite >2 years of active ART, and secondary NSV, defined as NSV in individuals who had previously achieved viral suppression before experiencing viral rebound. In Fig. 1 (and Fig. S1), we show the dynamics of viral load and CD4⁺ T-cell counts in representative primary and secondary NSV cases.

**Fig. 1.**
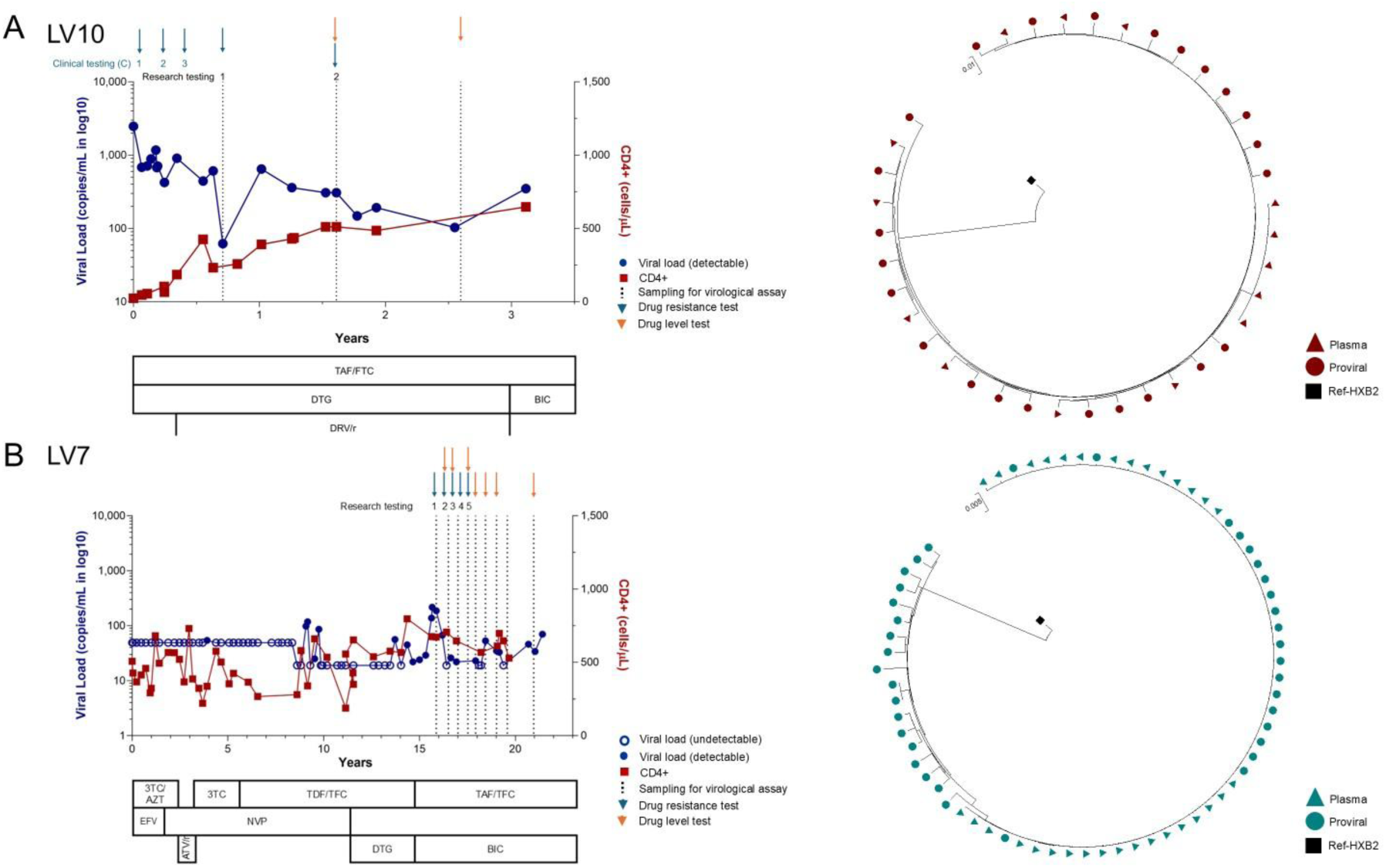
Representative (A) primary and (B) secondary NSV cases showing longitudinal clinical courses and phylogenetic trees. Longitudinal viral loads (blue) and CD4⁺ T-cell counts (red) are shown with antiretroviral regimens, resistance mutations, and drug level testing. Arrows indicate time points for research and clinical sampling. Phylogenetic trees on the right depict plasma and proviral HIV sequences. TAF, tenofovir alafenamide; FTC, emtricitabine; DTG, dolutegravir; DRV/r, darunavir/ritonavir; BIC, bictegravir; TDF, tenofovir disoproxil fumarate; ATV/r, atazanavir/ritonavir; RPV, rilpivirine; DRV/c, darunavir/cobicistat

To exclude the possibility of ART non-adherence, we measured antiretroviral drug concentrations in all participants. Most participants had intracellular levels of tenofovir and emtricitabine measured. Tenofovir levels reflect intermediate-term ART adherence (over the past ∼2 months) while emtricitabine levels reflect short-term adherence. Tenofovir and emtricitabine levels were assessed by dried blood spot in 12 participants (Fig. S2), and all exceeded thresholds consistent with ≥85% intermediate and short-term ART adherence (1,850 pmol/punch for tenofovir and 2.5 pmol/punch for emtricitabine) (*10*). Tenofovir and emtricitabine levels did not differ significantly between the NSV cohort and ART-suppressed historical controls (*1*). Dolutegravir and darunavir plasma levels were assessed in two participants, and both were consistent with adherence to ART (Table S2). Cabotegravir and rilpivirine levels were measured in one participant, and both exceeded four times the protein-adjusted concentrations required for 90% inhibition of viral replication (664 ng/mL and 50 ng/mL, respectively) (*11*). Collectively, these results highlight that NSV in our cohort were not clearly attributable to either drug resistance (based on GSS≥2, Table S1) or ART non-adherence.

### Clinical features of primary and secondary NSV

Of the three participants with primary NSV, two were diagnosed as adults in the setting of advanced HIV (nadir CD4 count <25 cells/mm^3^ for both) and both had very slow viral load decay after ART initiation without viral load suppression for >3 years (Fig 1, Fig. S1). The other participant was diagnosed as an infant with perinatal infection. ART initiation occurred ∼1 month after delivery, but the clinical course has been complicated by prolonged viral load decay and persistent viremia for >20 years despite multiple changes to the ART regimen and initiation of cabotegravir/rilpivirine long-acting ART. This individual had 13 HIV drug resistance tests performed, with no evidence of drug resistance to any of the contemporary regimens with GSS≥2 at all time points (Figure S1). Her CD4 count was appropriately high as a child and has remained stable as an adult. Furthermore, the NSV has persisted despite switching to long-acting injectable antiretroviral drugs (cabotegravir/rilpivirine) with on-time injections.

Participants with primary NSV had a lower CD4⁺ T-cell nadir than those with secondary NSV (median 22 vs. 242 cells/mm^3^; Mann-Whitney *P*=0.02; Fig. S3), excluding one participant with perinatal HIV infection, as infants have dramatically higher CD4 counts compared to adults. Taken together, the distinctive clinical picture, characterized by persistent lack of viral suppression after ART initiation and a markedly lower CD4⁺ T-cell nadir (in the adult participants), suggested potential differences in the HIV reservoir. We hypothesized that primary NSV would display distinct reservoir and phylogenetic characteristics compared with secondary NSV.

### HIV reservoir quantification in primary and secondary NSV

The levels of intact and defective proviruses were assessed by intact proviral DNA assay (IPDA). All three primary NSV participants had a dramatically elevated percentage of intact proviruses that was significantly higher than either secondary NSV or ART suppressed individuals (primary vs secondary, median 91% vs 10%, P=0.007; primary vs ART suppressed, median 91% vs 9%, P=0.0004, Fig. 2A). Primary NSV participants were found to have significantly larger intact proviral reservoirs compared to those with secondary NSV (median 3.7 vs. 2.2 log_10_ intact HIV DNA copies/million CD4 cells; *P*<0.001; Fig. 2B) or ART suppressed individuals (median 3.7 vs 1.6 log_10_ intact HIV DNA copies/million CD4 cells). There were no significant differences in defective or total HIV DNA size between primary and secondary NSV participants, and both primary and secondary NSV participants demonstrated larger intact, defective, and total reservoir size compared to ART suppressed individuals (Fig. 2B-D). Longitudinal IPDA measurements were available for 9 participants (3 primary and 6 secondary NSV) across a median of 39 weeks. There was no significant change in reservoir size over time for either primary or secondary NSV participants (Fig. S4).

**Fig. 2.**
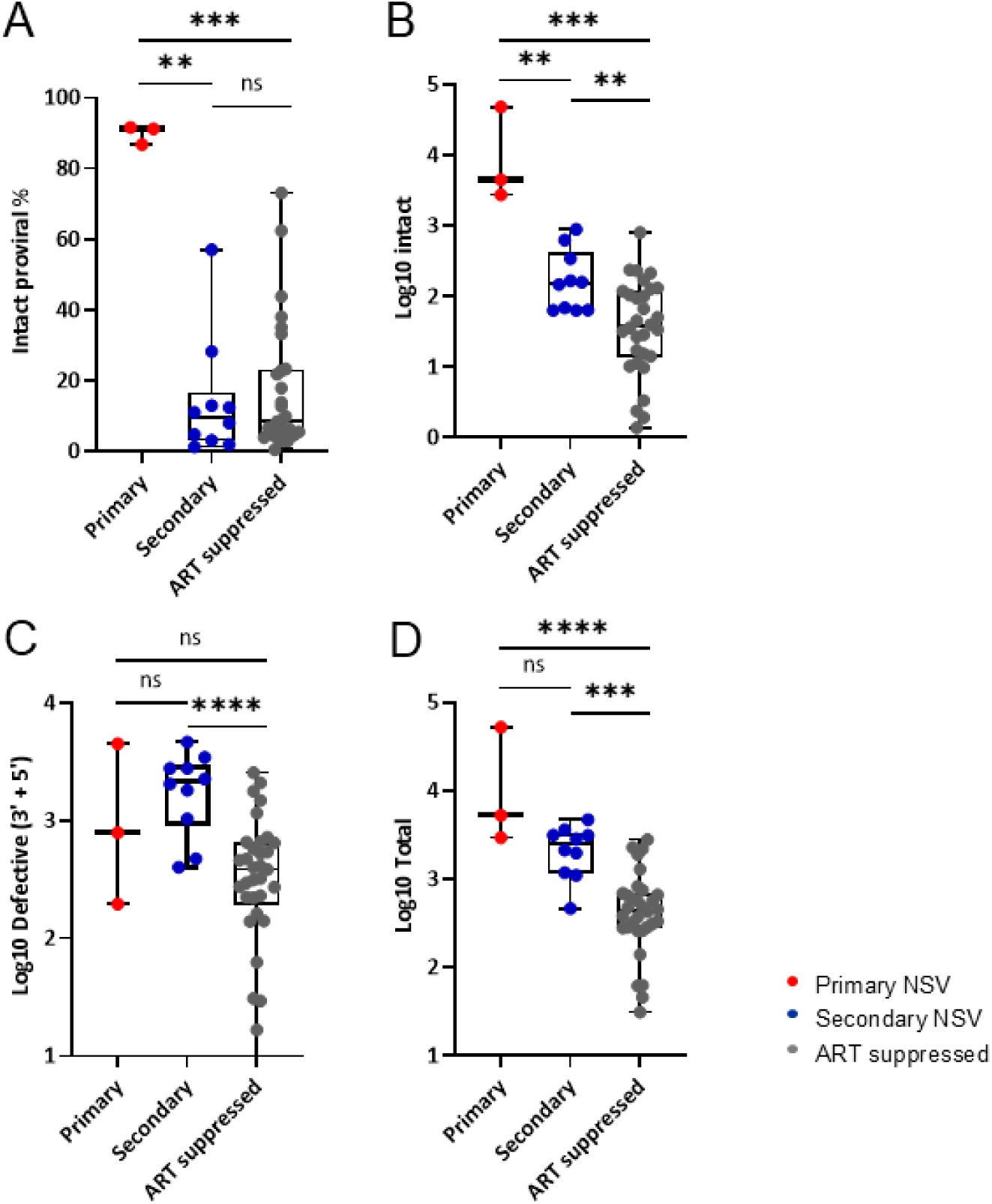
Intact proviral DNA assay showing total, intact, and defective HIV proviral DNA levels. (A) Proportion of intact proviruses among total proviruses in PBMCs from individuals with primary (red) and secondary (blue) NSV as measured by intact proviral DNA assay. (B–D) Levels of intact, defective, and total proviruses per million PBMCs. Horizontal bars indicate medians, and whiskers represent the full data range (minimum to maximum). Comparisons were performed using the Mann–Whitney *U* test.

### Proviral and plasma HIV RNA sequence signatures of primary versus secondary NSV

Using limiting dilution single-genome amplification, we obtained a total of 2,612 near-full length single-genome proviral sequences and 429 single-genome plasma sequences (*pol-env* or *pol*) for the 15 NSV participants (a median of 38 proviral sequences and 20 plasma HIV sequences analyzed per participant). We had previously reported that the plasma sequence of secondary NSV participants were largely present as part of 1 or 2 large clones (*1*). Neighbor-joining trees (NJTs) derived from plasma HIV sequences demonstrated that unlike secondary NSV participants, the plasma of primary NSV participants showed relatively few clones (Fig 3). Primary NSV participants had significantly fewer plasma sequences present as part of a clone compared to secondary participants (median 36% vs 87%, *P*=0.004; Fig. 4A). In conjunction with the IPDA data, the results suggest that while secondary NSV is driven by one or two large, transcriptionally active HIV-infected CD4 clones, primary NSV results from a far broader degree of proviral expression in the setting of a significantly larger reservoir.

**Fig. 3.**
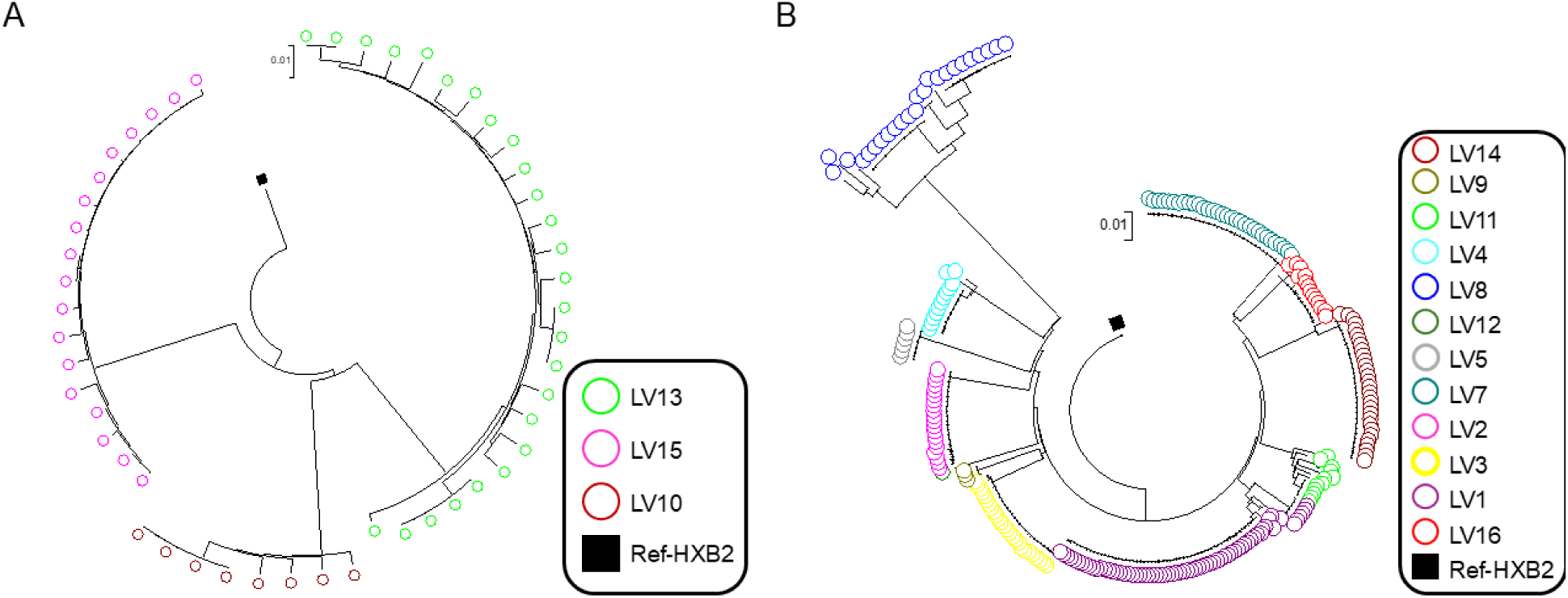
Phylogenetic relationships of plasma-derived HIV sequences from primary and secondary NSV. Neighbor joining trees constructed from plasma HIV sequences of individuals with (A) primary and (B) secondary NSV. Each color represents sequences from an individual participant. Primary NSV samples showed largely non-clonal sequences, whereas secondary NSV exhibited multiple clusters of identical or near-identical sequences.

**Fig. 4.**
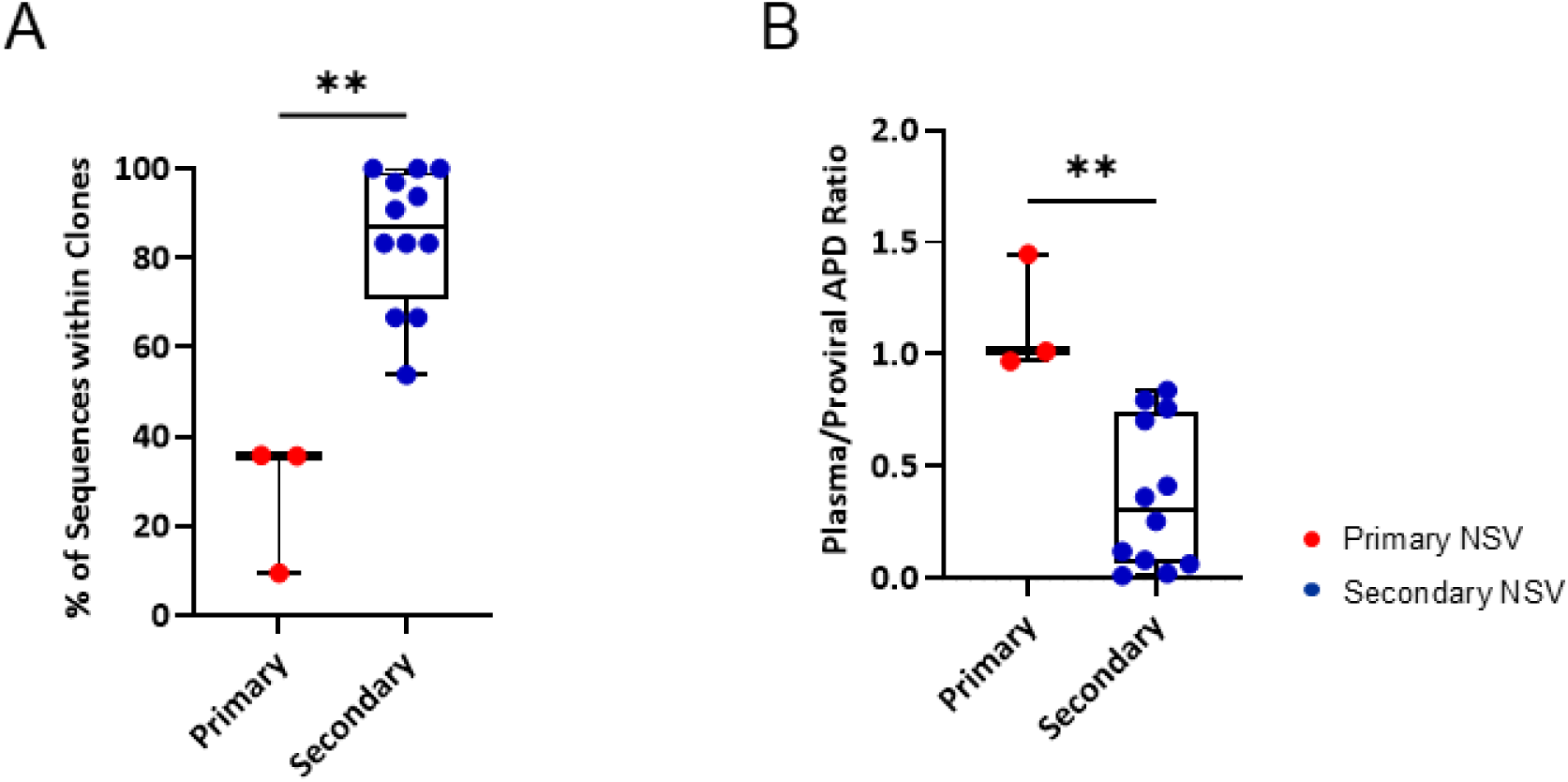
Clonal lineage composition and plasma/proviral diversity relationships in primary and secondary NSV. (A) Proportion of plasma sequences that belonged to expanded clonal lineages in primary (red) and secondary (blue) NSV. Secondary NSV cases showed a significantly higher fraction of clonally expanded sequences compared with primary NSV. (B) Ratios of plasma to proviral average pairwise distance, reflecting the relative diversity of circulating versus reservoir viruses. Horizontal bars indicate medians, and whiskers represent the full data range (minimum to maximum). Statistical comparisons were performed using the Mann–Whitney *U* test.

Additionally, we assessed viral diversity using average pairwise distance (APD) and viral evolution by root-to-tip distance. We did not detect any significant differences between primary and secondary NSV participants in the level of viral diversity of plasma and proviral intact sequences (Fig. S5A–B). Interestingly, the ratio of plasma HIV RNA to proviral DNA diversity was significantly higher in primary NSV (median APD ratio, primary vs secondary: 1.0 vs 0.3; P = 0.004; Fig. 4B). These finding highlights that the plasma sequences in secondary NSV represent a minor subset of the viral reservoir while in primary NSV, a far larger fraction of the reservoir is reactivated and contributing to the plasma viremia. The extent of viral evolution by root-to-tip distance measured on plasma or proviral sequences also did not differ between primary and secondary NSV groups. In those participants with longitudinal measurements, we detected no signs of viral evolution and active viral replication (Fig. S5C–D) and we found no significant differences in plasma sequences by the panmixia test (Figure S6). Together, these data show the lack of viral evolution over time for both the primary and secondary NSV groups.

### HIV phenotypic signatures in primary and secondary NSV

Full-length *env* clones were used to generate reporter pseudoviruses and assessed for their ability to facilitate entry into cells expressing a low surface density of CD4 (*12, 13*), a proxy for macrophage tropism (*14*). HIV envelopes from both primary and secondary NSV participants were able to efficiently infect cells with high CD4 expression but were highly inefficient in infecting cells with low CD4 expression (Fig. S7) (*12–14*), consistent with NSV in these participants being produced by CD4+ T cells and not macrophages.

### HIV sequence signatures in primary NSV

Among the primary NSV participants, we observed that within both the proviral and plasma sequences, deletions in the *vif-vpr* accessory protein region were common, with a range of deletion sizes that were rarely clonal (Fig. 5). There was some variation between the three primary NSV participants, with LV15 having the most conserved deletion in *vif-vpr* with only 19% of sequences without the deletion. LV10 had the largest proportion of intact sequences with 90% of plasma sequences and 95% of all proviral sequences being intact. LV13 had the lowest proportion of intact sequences at only 20%. LV13 also had the most extensive regional deletion, spanning *vif-env*. Notably, none of the 12 secondary NSV participants harbored similar targeted deletions in the *vif-vpr* or accessory protein regions of either the plasma or proviral sequences (Fig. S8).

**Fig. 5.**
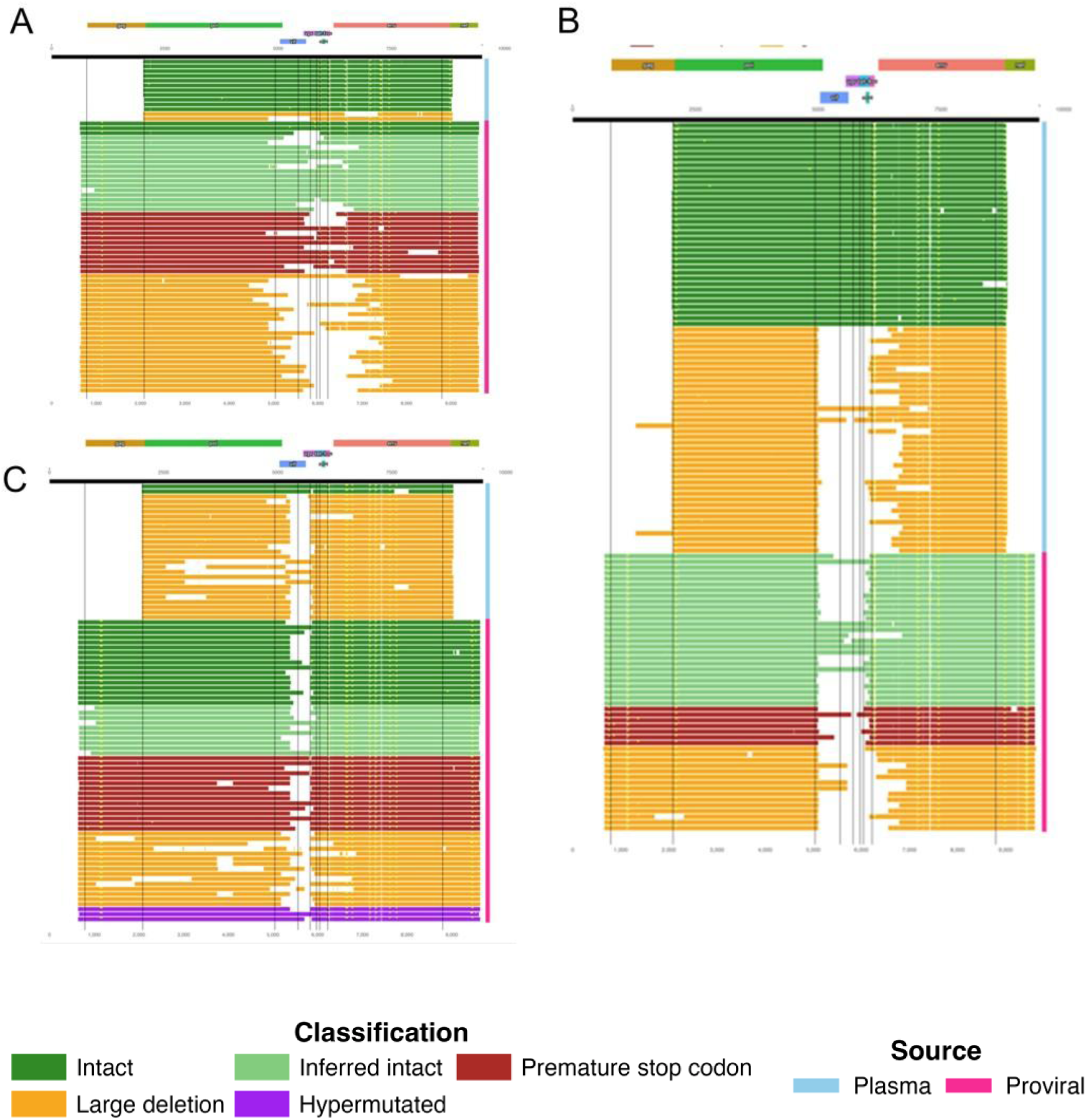
Proviral and plasma HIV structures and deletion patterns in primary NSV cases. Virograms from three primary NSV participants LV10 (A), LV15 (B), LV13 (C) showing intact genomes and internal deletions. Each row represents an individual proviral or plasma HIV genome aligned to the HXB2 reference. Note that all participants exhibited frequent *vif–vpr* deletions. Side bar shows sequence source plasma (blue) or proviral (pink). Color of bar indicates sequence intactness. Total length of bar shows sequenced region with plasma being either *pol-env* or *pol* and proviral sequences being near full length.

### HIV Transcriptional Landscape Analysis

We next used a multiplex droplet digital PCR assay to assess the intracellular HIV transcriptional landscape in those with primary and secondary NSV (Fig. 6A). We quantified levels of 6 different HIV RNA transcripts, including total initiated (TAR), 5’ elongated (R-U5-pre-Gag), mid-transcribed and unspliced (Pol), 3’ transcribed (Nef), completed (U3-polyA), and multiply spliced (Tat-Rev) HIV RNA. Individuals with primary NSV generally exhibited higher levels of intracellular HIV RNA transcripts, but only levels of HIV *pol* transcripts were significantly higher in primary compared to secondary NSV.

**Fig. 6.**
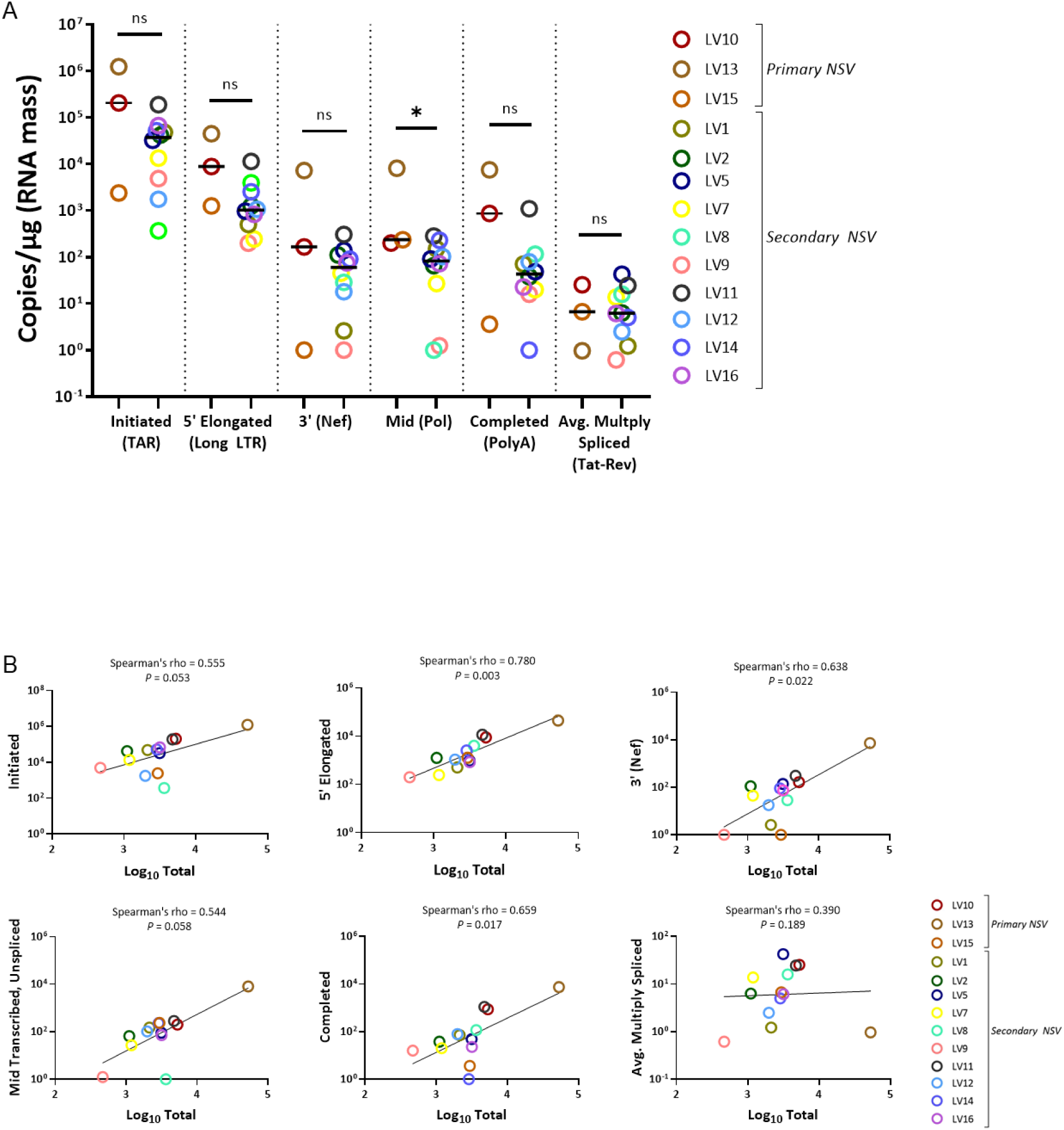
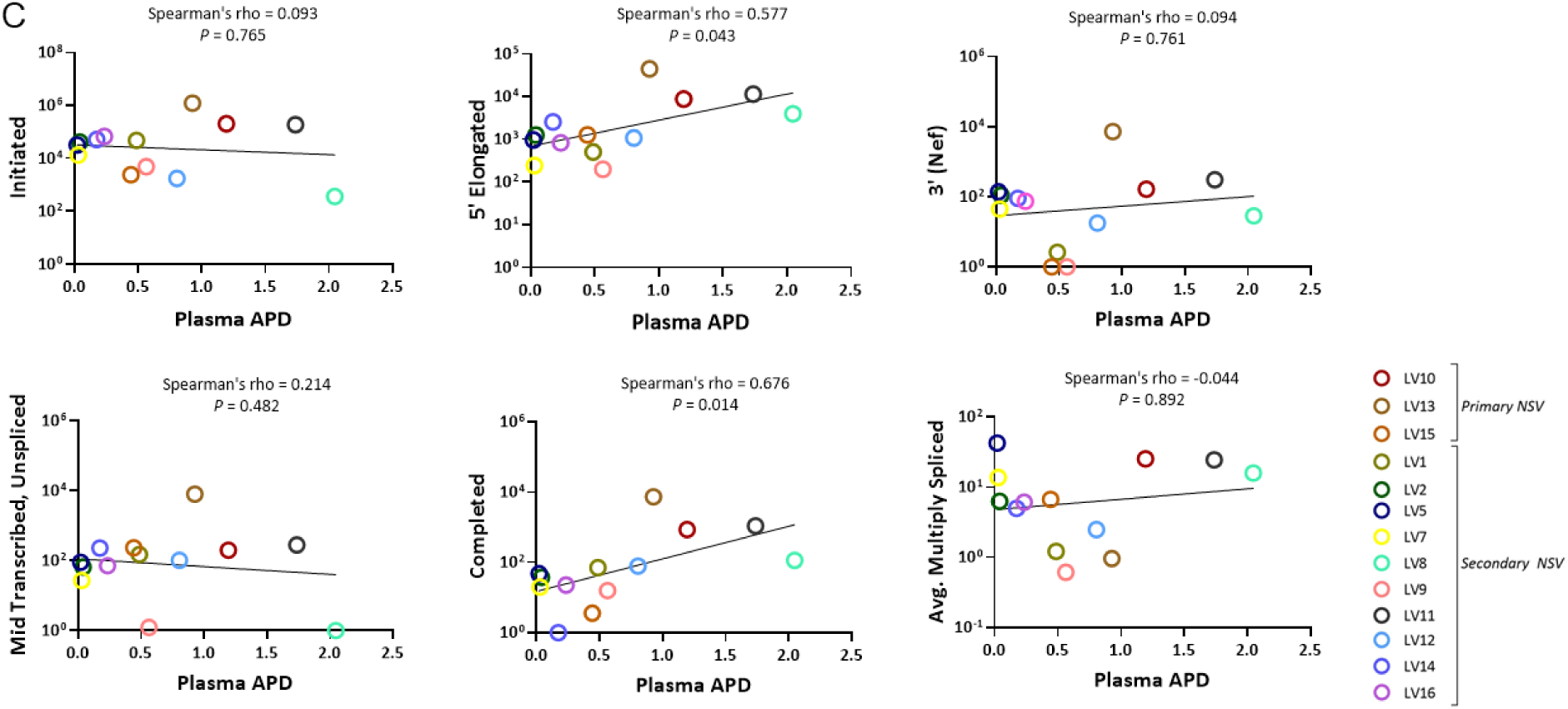
HIV transcriptional activity and its correlation with reservoir size and viral diversity. (A) Levels of different HIV RNA species measured in primary and secondary NSV participants. Although not statistically significant, primary NSV tended to show higher transcript levels, particularly in Mid (pol). (B) Correlation between total reservoir size (total HIV DNA levels by the IPDA in log_10_ HIV copies/million CD4 T cells) and HIV RNA transcript levels, showing that larger reservoirs were associated with greater transcriptional output. (C) Correlation between plasma average pairwise diversity and HIV RNA transcript levels, indicating that more diverse viral populations were linked to higher transcriptional activity. Horizontal bars represent medians; error bars represent min–max ranges. Comparisons were performed by Mann–Whitney *U* test and correlations were assessed using Spearman’s rank correlation coefficient (ρ).

We also examined how transcriptional activity was associated with reservoir size and plasma viral diversity. Total HIV DNA levels showed a positive correlation with most HIV transcripts except for multiply-spliced transcripts (Fig. 6B), with weaker associations of HIV transcripts with the intact reservoir (Fig. S9), suggesting that much of the intracellular HIV RNA is likely produced by defective proviruses. In addition, plasma viral diversity, measured by APD, also showed a positive association with later stages of HIV transcript levels, particularly in 5′ elongated and completed transcripts (Fig. 6C).

### Inflammatory Markers as a Surrogate for Systemic Inflammation

To assess whether systemic inflammation differed between primary and secondary NSV, we compared a panel of soluble inflammatory and immune activation markers across cohorts. Overall, levels of most cytokines and soluble markers were comparable between primary and secondary NSV participants. Compared to ART-suppressed individuals, primary NSV participants had higher levels of CRP while secondary NSV participants had higher levels of CD163 (Fig. S10). These inflammatory markers were not significantly different between primary and secondary NSV participants. These findings suggest that the primary and secondary NSV subtypes are not explained by broad differences in systemic inflammatory profiles.

## Discussion

Despite the effectiveness of modern ART regimens, some individuals continue to have persistent NSV regardless of appropriate ART regimens and adherence, resulting in a clinical and scientific dilemma. Most studies on NSV have focused on individuals who had initial plasma HIV suppression and then developed persistent low-level viremia (*1, 9, 15, 16*). In addition to this population (termed secondary NSV), we report another distinct subset of NSV, termed primary NSV. Primary NSV is defined as individuals who experienced a very prolonged period of viremia after ART initiation without virologic suppression, in one case with NSV for >20 years. We were able to identify distinct profiles of primary NSV participants relative to secondary NSV, including low CD4 nadir, an exceptionally large intact proviral reservoir, a more diverse composition of plasma viremia, and distinct deletions in accessory genes, highlighting potential mechanisms in persistent viremia.

Both primary and secondary NSV participants had excellent ART adherence by drug level testing and were receiving at least 2 active antiretroviral drugs. Drug level testing included intracellular levels of tenofovir and emtricitabine, which confirm both high levels of short– and intermediate-term ART adherence (*17, 18*). In both groups, we found no evidence for active viral replication or evolution, as there were no significant changes in either the size of the reservoir over time or in the phylogenetic root-to-tip distances of longitudinal viral sequences, a sign of ongoing viral evolution. These results demonstrate that insufficient ART-mediated viral suppression is unlikely to be the cause of either primary or secondary NSV. Therefore, strategies altering the ART regimen or adding new antiretroviral drugs (intensification) are unlikely to improve the control of NSV.

In addition to the clinical course of viremia after ART initiation, we were able to identify key viral reservoir and sequence features that distinguished those with primary versus secondary NSV and pointed to potential mechanisms that may differ between the two groups. While secondary NSV participants had a larger intact reservoir compared to ART-suppressed individuals, we found that primary NSV participants had exceptionally large intact reservoirs that were a median >1 log_10_ higher than those with secondary NSV and with an intact reservoir by the IPDA that comprised >80% of the total reservoir in each of the three cases. In two out of the three primary NSV cases, ART was initiated at a very low CD4 count, suggesting the widespread seeding of a depleted CD4 T cell population. The last primary NSV participant was infected perinatally and started ART approximately 1 month after delivery. The reservoir characteristics of this participant are in distinct contrast from previously reported reservoir features of perinatally infected infants with early and sustained ART (initiated within a median of 4 months and sustained for 20 years) that showed a significantly smaller intact HIV reservoir compared to ART-suppressed adults (*19–21*).

We previously reported that individuals with secondary NSV had plasma comprised of one or two large clonal lineage sequences that point to a hyperactive clonally expanded HIV reservoir as the source of the viremia. Interestingly, primary NSV participants demonstrated far more diversity in the plasma sequences that highlight the activation of a far broader swath of the HIV reservoir. This finding is also evident by our analysis demonstrating that plasma and proviral sequence diversity were comparable by average pairwise distance. In contrast, plasma HIV diversity was far lower than that of the intact proviral DNA in secondary NSV, demonstrating that only a small subset of the HIV reservoir was contributing to plasma viremia. These findings align with prior studies demonstrating that persistent plasma viremia in secondary NSV is frequently driven by clonally expanded infected cells (*1*). Our observations in primary NSV are consistent with a model in which plasma viremia arises from broader expression from a larger proportion of cells in the HIV reservoir, potentially involving numerous distinct proviral lineages rather than dominance by a single expanded clone (*22*). While broader reservoir contributions can be explained by heterogeneous populations of infected CD4⁺ T cells, they raise the possibility of additional cellular or tissue-based reservoirs that warrant further investigation (*23*).

The abundant primary NSV-specific *vif–vpr* deletion is particularly intriguing and has been described in a recent report of NSV that suggests that these viruses may arise from a macrophage reservoir (*24*). In contrast, the viruses from both the primary and secondary NSV participants in our cohort, including those with *vif-vpr* deletions were determined to be T cell tropic. Hariharan *et al.* has also described individuals with similar deletions (*25*), suggesting that superinfection of cells containing both wild-type and deleted mutants can facilitate the propagation of deleted genomes and contribute to NSV. It is not clear, though, why this phenomenon was only seen in our primary NSV participants, and further studies are warranted to elucidate how the *vif–vpr* deletion influences host immunity or viral fitness and thereby contributes to a distinct form of NSV. In addition, the identification of these deletions by near-full length sequencing highlight the limitations of the IPDA, which only evaluates a very limited portion of the HIV genome to determine intactness.

In summary, we report that NSV is comprised of two distinct subsets of individuals, including a novel group with primary NSV characterized by prolonged viremia after ART initiation, an exceptionally large intact reservoir and highly diverse plasma virus populations arising from transcriptionally active proviral reservoirs, and no evidence of ongoing evolution. Ultrasensitive HIV RNA viral load assays have shown that the vast majority of PWH have very low levels of residual viremia despite apparent suppression by commercial viral load tests (*26*). Thus, understanding the drivers and mechanisms of non-suppressible viremia has implications for understanding HIV reservoir persistence for all PWH and will inform efforts to design strategies for HIV reservoir clearance and control.

## Materials and Methods

### Participants

In a prospective observational study, we enrolled ART-treated participants with three or more HIV RNA levels over 40 copies/mL over 24 months. A participant enrolled in the HIV Eradication and Latency (HEAL) cohort, a biorepository of Brigham and Women’s Hospital, was also included. Samples were obtained from one or more time points, enabling longitudinal analysis in a subset of participants (Supplementary Figure 1). Post-treatment noncontrollers from prior AIDS Clinical Trials Group (ACTG) analytical treatment interruption studies were included as a control group for IPDA analyses (*27*), and chronically treated, ART-suppressed participants from the A5345 study were included as a control group for inflammatory marker analyses (*28*).

### Antiretroviral drug level testing

For plasma antiretroviral drug level testing, darunavir, dolutegravir, cabotegravir, and rilpivirine concentrations were quantified using liquid chromatography coupled with tandem mass spectrometry (LC–MS/MS). For dried blood spot (DBS) testing, 25 μl aliquots of whole blood were applied in quintuplicate onto Whatman 903 protein saver cards, as previously described (*18, 29*). The cards were dried at room temperature for at least 3 hours (up to overnight) before storage at −80 °C until analysis. Intracellular tenofovir diphosphate and emtricitabine triphosphate were measured from two 7-mm punches extracted with 2 ml of methanol: water to generate a lysed cellular matrix, using a validated method that was subsequently adapted for tenofovir alafenamide–containing regimens. The assay demonstrated linearity from 25 to 6,000 pmol per sample for tenofovir and from 0.1 to 200 pmol per sample for emtricitabine (*18, 30*).

### DNA isolation and HIV reservoir quantification

Genomic DNA was isolated from PBMCs with the QIAamp DNA Micro Kit (Qiagen, Hilden, Germany; catalog no. 56304) and quantified using a Nanodrop spectrophotometer (Applied Biosystems, Thermo Fisher). To approximate reservoir size, near–full-length proviral sequences were subsequently analyzed as outlined below. In addition, intact and defective proviruses were quantified using the IPDA performed by Accelevir Diagnostics as previously described (*31*), and expressed as log₁₀ copies per 10⁶ CD4⁺ T cells.

### Near-full-length proviral sequencing, sequence alignments, quality control and neighbor joining analyses

Extracted DNA was subjected to near full-length sequencing (NFL-seq) in limiting dilution, as previously described (*32*). Sequences were classified as intact or assigned to categories of defective proviruses (e.g., 5′ defects, deletions, hypermutations, or inversions) using a published proviral intactness pipeline (*30*). In brief, after alignment to HXB2, sequences were designated as carrying large deleterious defects if they exhibited <8,000-bp amplicon size, out-of-frame indels, premature or lethal stop codons, internal inversions, or ≥15-bp packaging signal deletions. Hypermutations associated with APOBEC3G/3F were identified using the Hypermut 2 program (Los Alamos National Laboratory HIV Sequence Database). Sequences lacking these defects were categorized as “genome-intact.” Multiple sequence alignment was performed with MAFFT v7.2.0, and neighbor-joining trees were inferred using MEGA 6.0.

### Plasma HIV RNA sequencing

Plasma HIV RNA was extracted as previously described (*33*). Extracted RNA was endpoint-diluted to achieve single-genome sequencing conditions, defined as no more than one template per well and ≤25% of wells positive for HIV. Primers targeting a 6.7-kb *pol–env* region were used (Supplementary Data 1). Amplification and sequencing were performed as previously described (*1*). Plasma sequences differing by no more than two nucleotides from NFL proviral sequences were classified as part of the same clonal cluster.

### Resistance mutation and GSS

For each sequence, the genotypic susceptibility score (GSS) relative to the participant’s current ART regimen was calculated using the Stanford HIV Drug Resistance Database scoring system (*34*). This algorithm assigns weighted penalty scores to each resistance mutation–drug combination, ranging from 0 (no expected effect) to 60 (high-level resistance). For every antiretroviral drug, the cumulative penalty score was obtained by summing the values for all resistance mutations present. The corresponding GSS values were then defined as follows: 1 (penalty score 0–9; susceptible), 0.75 (10–14; potential low-level resistance), 0.5 (15–29; low-level resistance), 0.25 (30–59; intermediate resistance), and 0 (≥60; high-level resistance). The overall sequence GSS was calculated as the sum of the GSS values for each drug included in the participant’s regimen.

### Phylogenetic and diversity analyses

Average pairwise distance (APD) was used to quantify intrapatient sequence diversity. Sequences were aligned using MAFFT v7.2.0, and APD was calculated in MEGA version 6.0 (Molecular Evolutionary Genetics Analysis; Sudhir Kumar, Temple University, Philadelphia, PA) as the mean number of nucleotide differences per site across all pairwise comparisons, using the p-distance model with complete deletion of gaps.

Root-to-tip (RTT) distances were assessed using a series of tools from the Los Alamos National Laboratory (LANL) HIV Sequence Database (Los Alamos, NM) (*35*). Briefly, nucleotide substitution models were selected with FindModel, maximum-likelihood phylogenetic trees were constructed with TreeMaker, and RTT distances were calculated from the resulting Newick trees using the TreeRate tool. For all analyses, the root was fixed at the designated node 1 rather than node 0. RTT distance for each sequence was defined as the cumulative branch length from the root to the terminal node. The median RTT distance across all intact sequences from each individual was used as the representative value.

Based on the NJT generated in MEGA version 6.0 sequences that were branched together were isolated. They were evaluated using the Highlighter tool from the Los Alamos National Laboratory (LANL) HIV Sequence Database (Los Alamos, NM) (*35*). Sequences with ≤2 mismatches from the master sequence were considered clones of the master sequence. The number of intact plasma sequences in a clone was divided by the total number of plasma sequences to determine the percentage of plasma sequences in a clone for each participant. Panmixia analyses were performed using the *panmixia.py* script available in Michael Bale’s python_stuff GitHub repository (https://github.com/michaelbale/python_stuff). For each participant, proviral and plasma HIV nucleotide sequences were aligned to the HXB2 reference using MAFFT and trimmed to the common region (*pol–env, 2065-9010bp*). A two-column tab-delimited file mapping each sequence ID to its compartment (proviral or plasma) was generated. The *panmixia.py* pipeline calculates pairwise genetic distances between all sequences and partitions them into within– and between-group comparisons to derive the Kst statistic, which quantifies genetic differentiation between compartments (higher Kst values indicate greater structure). Statistical significance was assessed through 100,000 random permutations of group labels to generate a null distribution of Kst values. Kst and –log10(*p*) values were visualized for each participant to compare the magnitude and significance of compartmentalization. All analyses were restricted to alignments of equal length.

### Assessment of cellular tropism (T-tropic or M-tropic)

HIV Env-mediated entry was assessed using a previously described low-CD4 entry assay to infer viral cellular tropism (*12–14*). Briefly, full-length env genes were amplified from plasma HIV RNA, cloned into expression vectors, and used to generate luciferase reporter pseudoviruses. Entry efficiency was measured in Affinofile cells induced to express low or high levels of CD4, serving as proxies for adaptation to enter macrophage (M-tropism; able to efficiently enter cells expressing a low surface density of CD4) or CD4⁺ T cells (T-tropism; requires a high surface density of CD4 for efficient entry).

### Virogram Plot

Proviral fragments and insertion coordinates were derived from a pre-computed multiple-sequence alignment in FASTA format using a custom Python workflow (Biopython SeqIO). The first record was treated as the HXB2 reference; alignment columns were mapped to HXB2 nucleotide coordinates by incrementing reference positions only at non-gap sites (reference gaps skipped). For each sample sequence, contiguous stretches of alignment columns where both the reference and sample were non-gapped were recorded as fragment intervals in HXB2 coordinates. Putative insertion sites were defined as columns where the reference contained a gap and the sample contained a nucleotide; each insertion was anchored to the nearest flanking HXB2 coordinate (left if available, otherwise right). Sequence identifiers were parsed and normalized into fragment classes (Intact, Inferred intact, Premature stop codon, 5′ defect, Large deletion, Hypermutated). Virograms were generated in R (ggplot2/dplyr/forcats/patchwork/shadowtext/stringr): annotated HXB2 ORFs (gag, pol, vif, vpr, tat, rev, vpu, env, nef) were plotted at fixed coordinates in a faceted 3-frame panel above the fragment tracks; per-sample fragments were rendered as horizontal rectangles across HXB2 positions and colored by class. Insertion sites were clustered within 3 bp per sample and marked at the cluster mean position. Specimen origin (Proviral/Plasma), when available from an external sample metadata table, was indicated by a short colored “Source” bar to the right of the genome window.

### HIV Transcriptional Landscape Analysis

HIV transcripts, including total initiated (TAR), 5’ elongated (R-U5-pre-Gag), mid-transcribed and unspliced (Pol), 3’ transcribed (Nef), completed (U3-polyA), and multiply spliced (Tat-Rev) HIV RNA, were measured by RT-ddPCR. Primers/probes and reaction conditions were as described previously (*36, 37*). Each transcript was measured in at least 2 duplicate wells. HIV RNA copies were normalized to total cellular transcription (1µg of RNA, corresponding to about 10^6^ cells) according to the RNA concentration, the volumes used in the RT, and the fraction going into each ddPCR well.

### Inflammatory soluble marker levels

Cryopreserved plasma samples were thawed and analyzed in batch to assess inflammatory cytokines. Assays were run on 96-well plates. Duplicates were performed for 25% of the samples that were run on each plate. Mesoscale Discovery single and multiplex assays were performed according to instructions provided in kits for assessment of IFN-γ, IL-6, IL-8, IL-10, IP-10 (K15067M-1), TGF-β1-3 (K15241K-1), TNF-RI, TNF-RII (K151AEM-1), sCD163 (F21J4-3), FABP2 (F21M4-3), C-reactive protein (K151STD-1), MMP-9 (F21AMN-3), and sCD14 (K151T1R-2). The Mesoscale assays were analyzed with a MESO QuickPlex SQ 120MM Reader. ART suppressed comparators included participants from the ACTG (Advancing Clinical Therapeutics Globally for HIV/AIDS and Other Infections) A5345 study.

### Statistics

We analyzed the data using Mann–Whitney U tests (two-tailed), Fisher’s exact tests, and Wilcoxon’s signed-rank tests, as appropriate, unless otherwise specified. Correlations were assessed using Spearman’s rank correlation coefficient (ρ). No correction for multiple comparisons was performed in other analyses, which were considered exploratory. Adjustments for multiple comparisons were applied to the analyses of the number of HLA escape mutations per HIV gene. Statistical analyses were conducted using Prism (GraphPad Software, version 10, San Diego, CA).

### Study approval

The study protocol was reviewed and approved by the Mass General Brigham Institutional Review Board in accordance with the principles of the Declaration of Helsinki. Written informed consent was obtained from all participants prior to study procedures.

### Data availability

A portion of the secondary NSV sequence data was previously submitted to Genbank (Bio-Project: PRJNA973660).

## Acknowledgements

We are grateful for the contributions of the participants who made this study possible. We thank the investigators and study staff of the ACTG A5345 study team. We appreciate the support of the staff at the MGH sequencing core facility. We would like to acknowledge additional laboratory staff who processed the participant samples.

## Author contributions

J.Z.L. and B.E. Conceived, designed, and supervised the project; L.H, L.C, H.J, D.P.W, N.K, J.M.J, A.N.T, S.D, and J.M.L. participated in sample collection; T.A. performed sample processing; T.A, CK.K, A.M, S.C, E.A.D, G.E.E, D.D, A.A.S, J.K, and B.E. performed the experiments; T.A, CK.K, and F.M. performed the statistical analysis; T.A, CK.K, A.M, F.M, M.M, J.R.C.M, C.F, P.L.A, S.Z, S.B.J, S.S, S.Y, B.E, and J.Z.L. participated in data collection and analysis; T.A, CK.K, F.M, J.Z.L. wrote the original draft of the paper; all authors contributed to the final review of the paper and the editing.

