## Supplemental figures and tables for "Reservoir and Phylogenetic Signatures Identify Distinct Subsets of HIV-1 Nonsuppressible Viremia"

**SUPPLEMENTAL FIGURES**
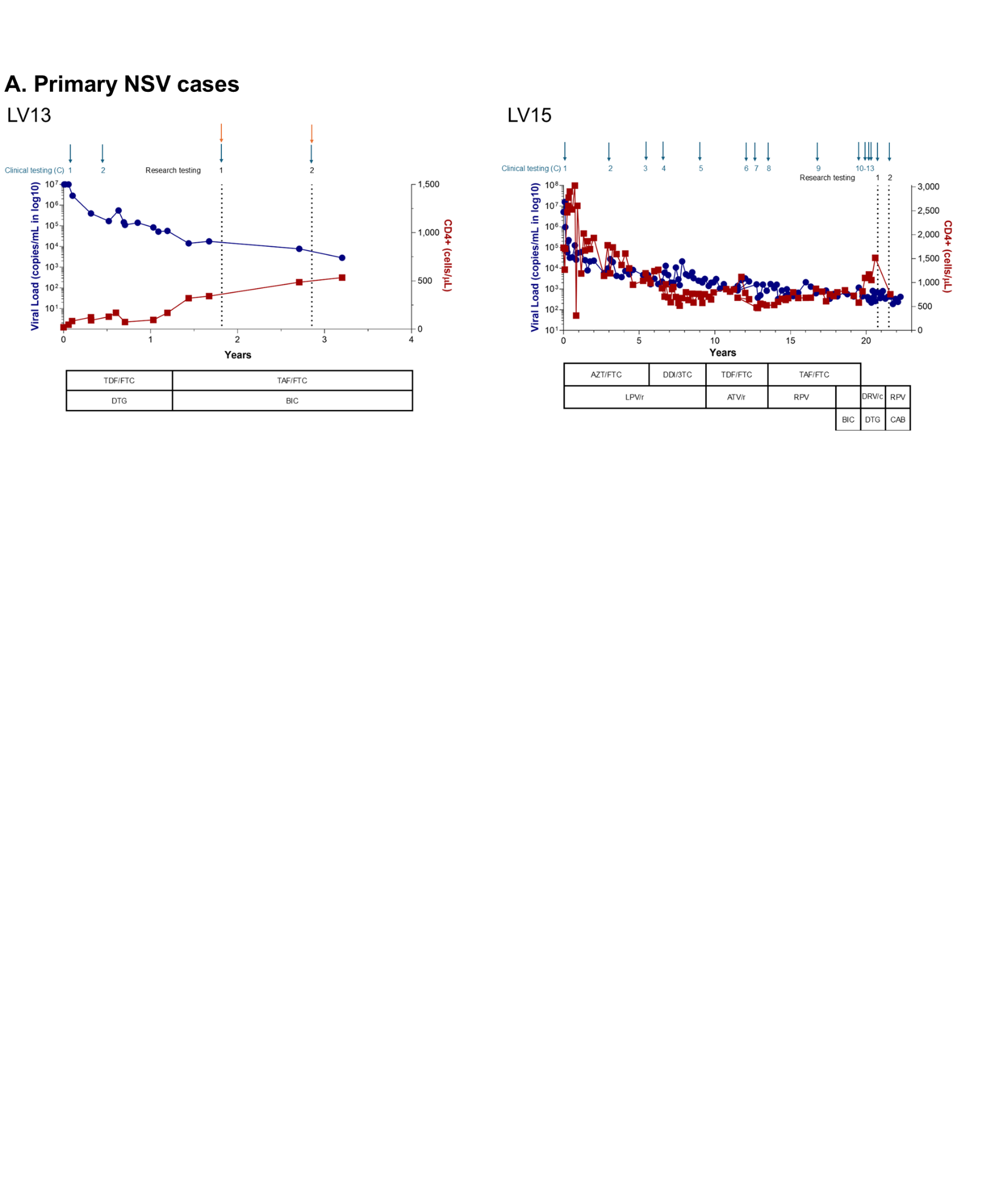

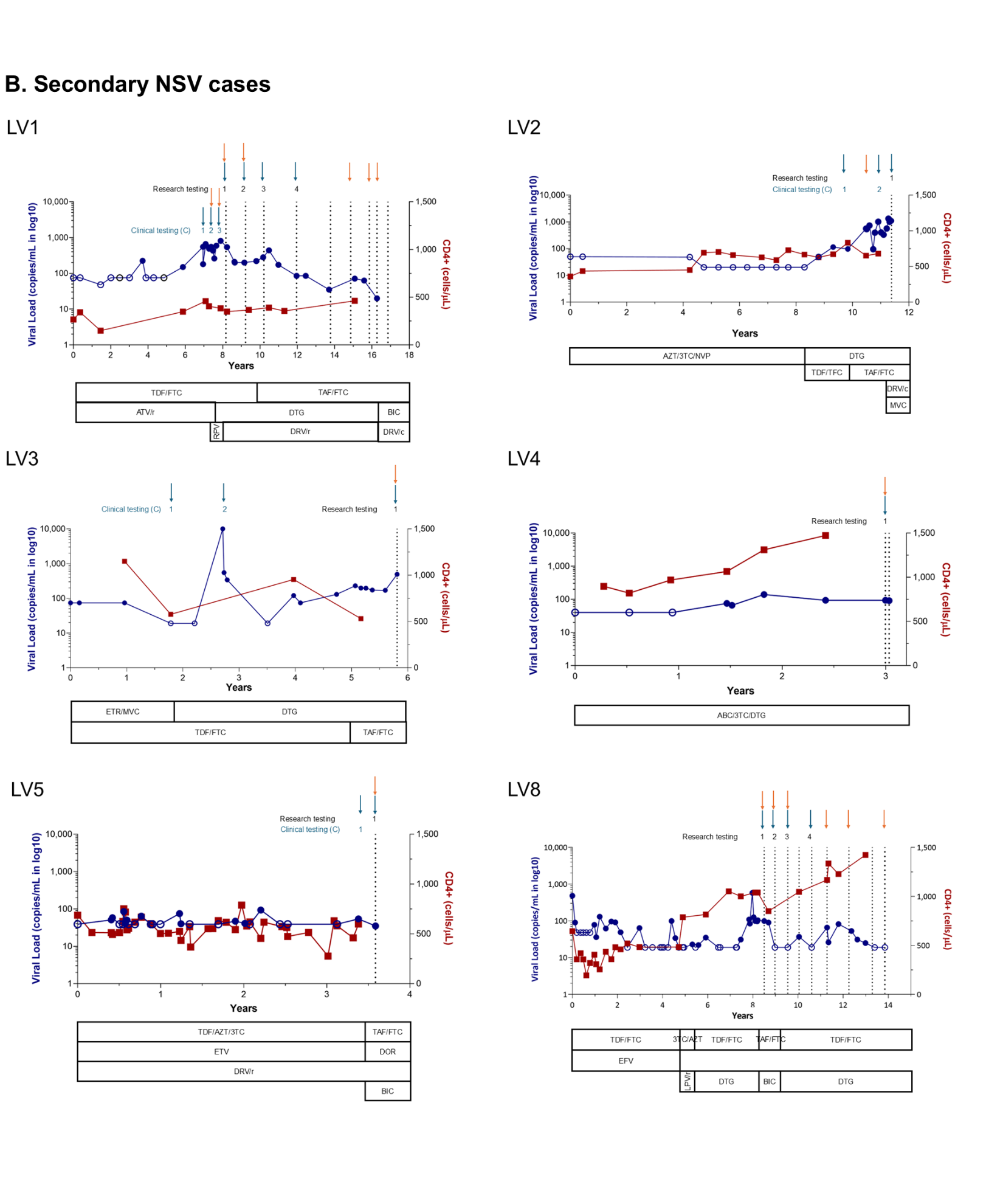

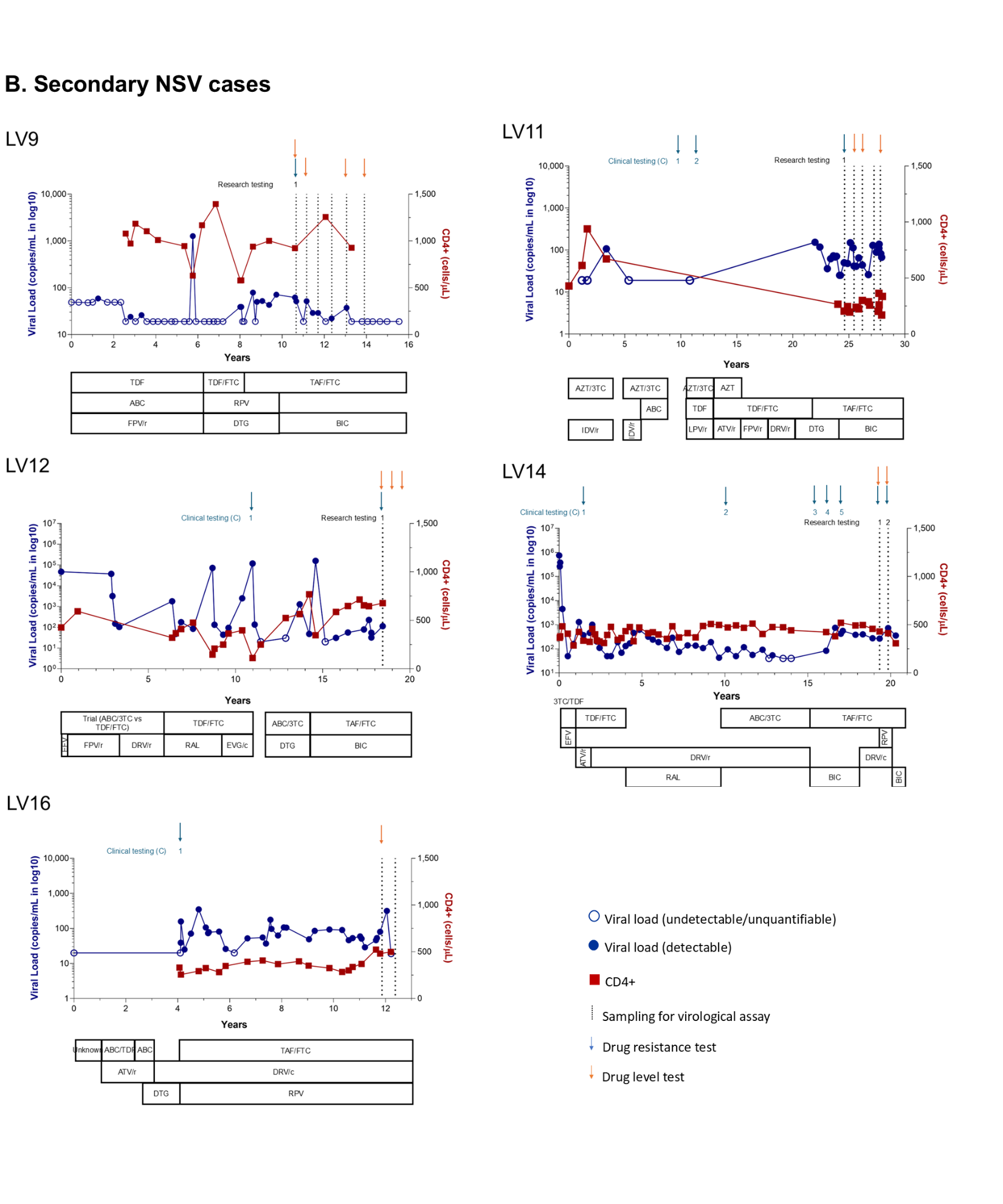

**Supplemental Figure 1**. CD4 count and viral load graphs for additional participants not shown in Figure 1, categorized by primary (A) or secondary (B) NSV.

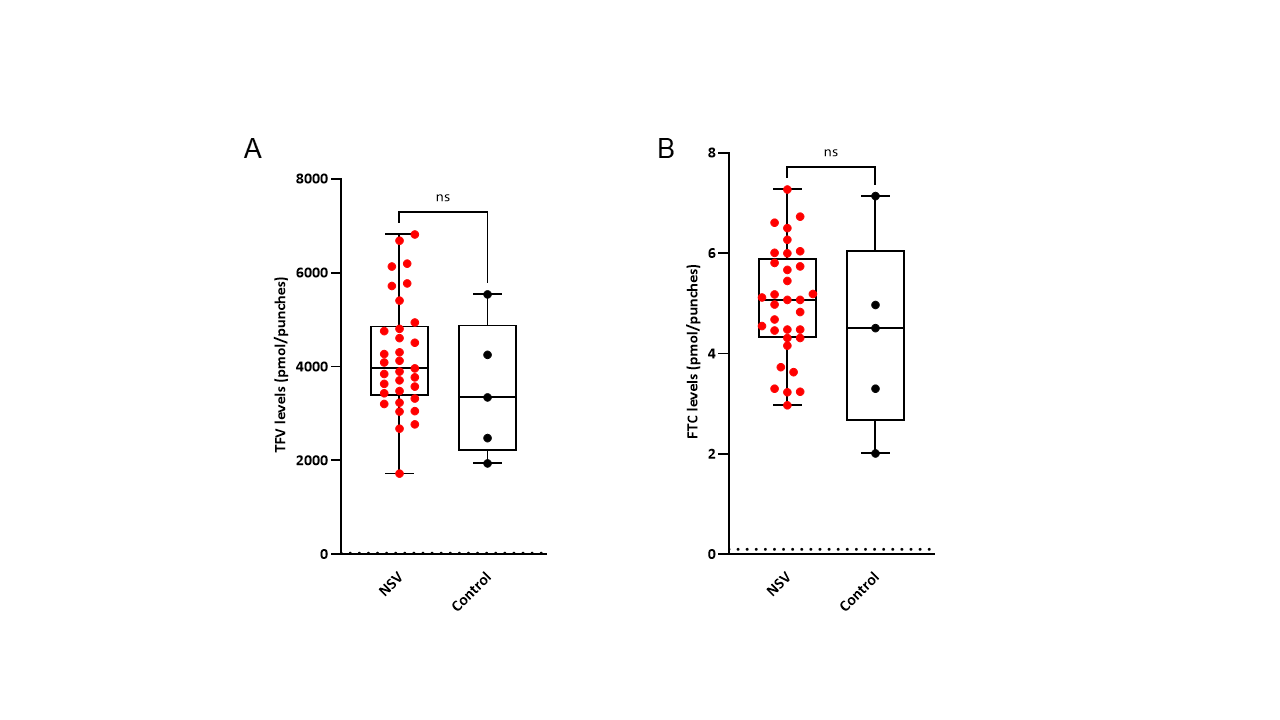

**Supplemental Figure 2.** Tenofovir (TFV) (A), and emtricitabine (FTC) (B) drug levels of non-suppressible viremia (NSV) participants for available all timepoints and ART suppressed control group. Comparisons were performed by Mann–Whitney *U* test.

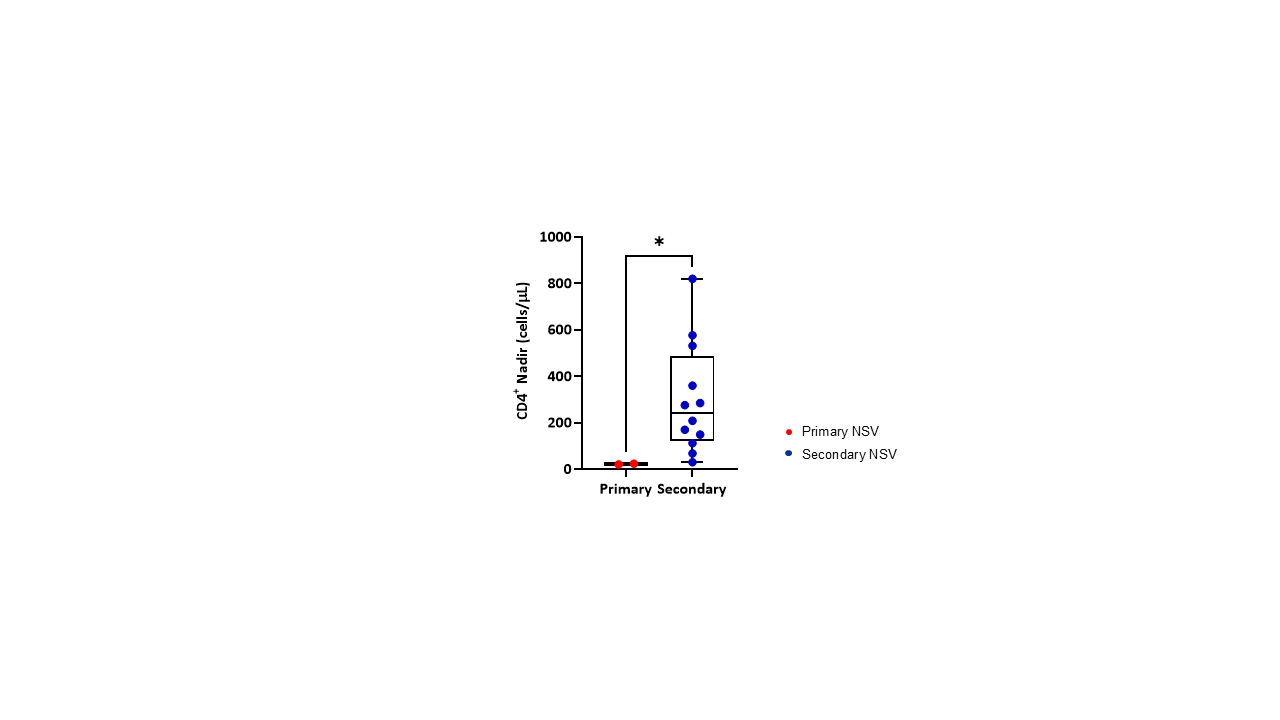

**Supplemental Figure 3.** CD4 nadir of primary versus secondary NSV participants. LV15 was excluded as infants have substantially different CD4 counts compared to adults. Comparisons were performed by Mann–Whitney *U* test. *p<0.05.

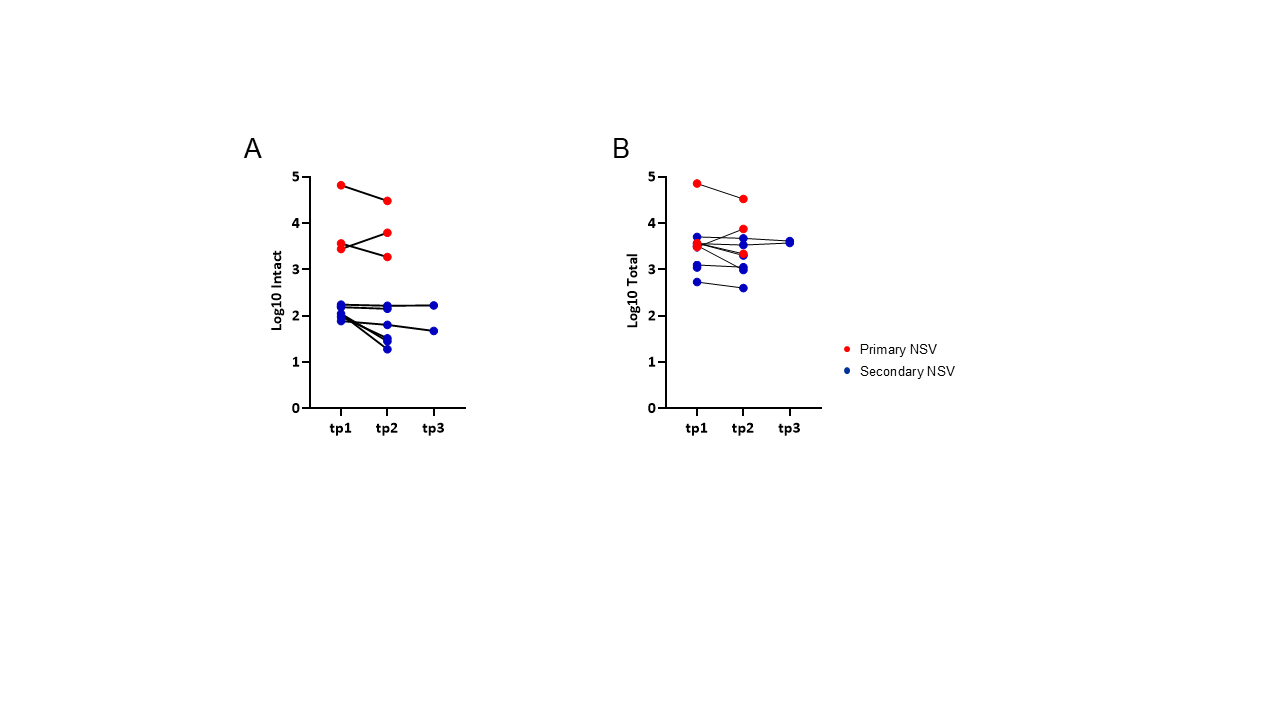

**Supplemental Figure 4.** Longitudinal levels of intact (A) and total (B) HIV reservoir size (log_10_ HIV DNA copies/million CD4 cells) by the IPDA assay for primary and secondary NSV participants with available longitudinal sampling.

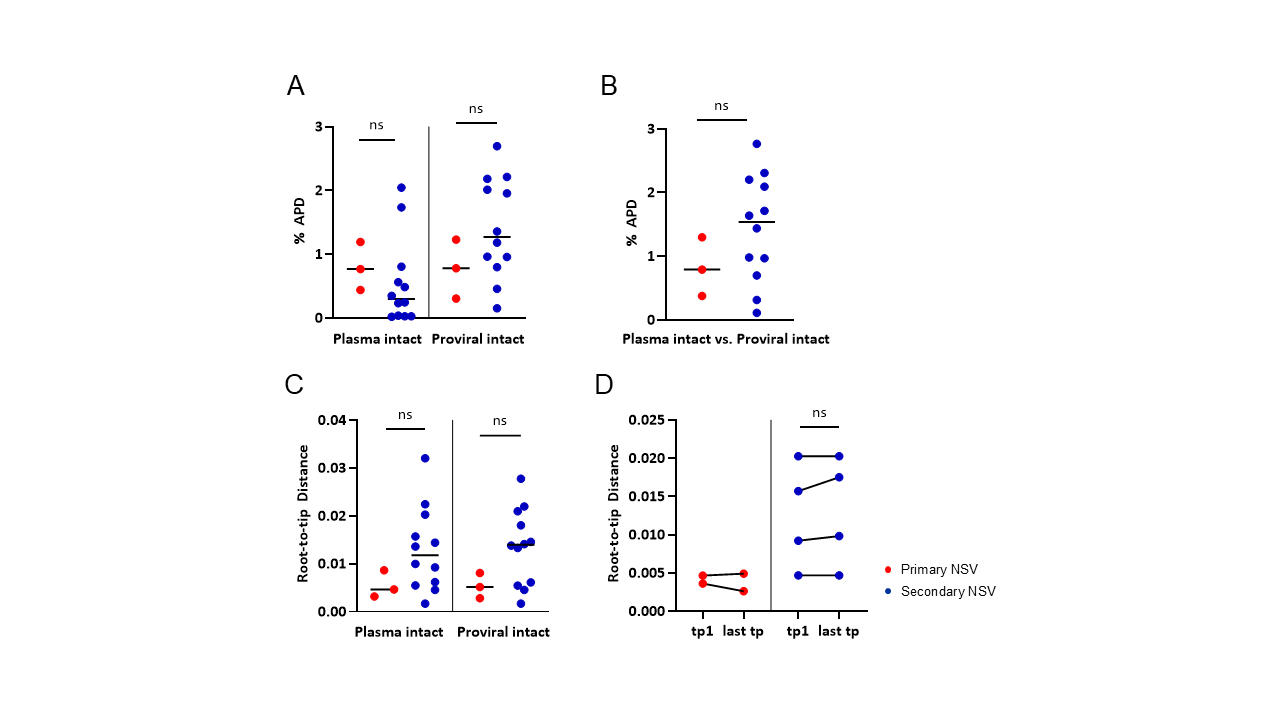

**Supplemental Figure 5.** Average Pairwise Distance within (A) or between (B) plasma and proviral intact groups, and cross-sectional (C) or longitudinal (D) Root to Tip distance of primary and secondary NSV participants. Comparisons were performed by Mann–Whitney *U* test.

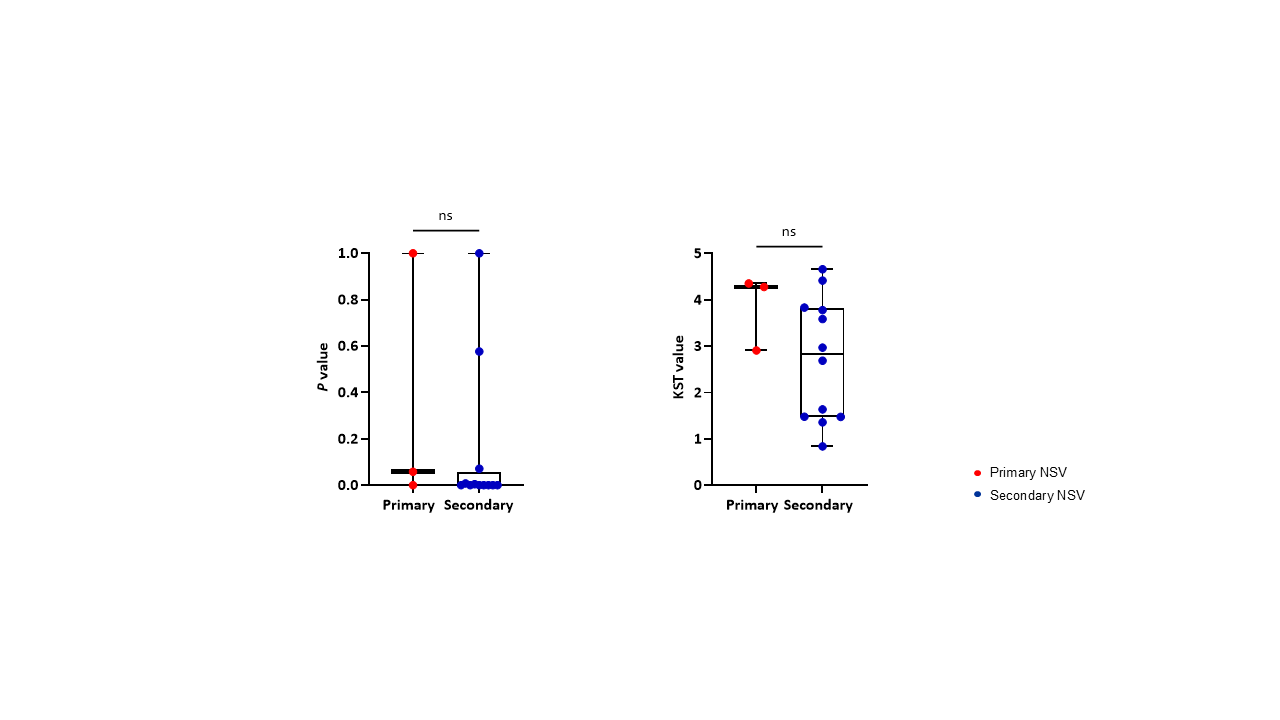

**Supplemental Figure 6.** Panmixia of primary and secondary NSV participants. No significant correlation between the two groups. Comparisons were performed by Mann–Whitney *U* test.

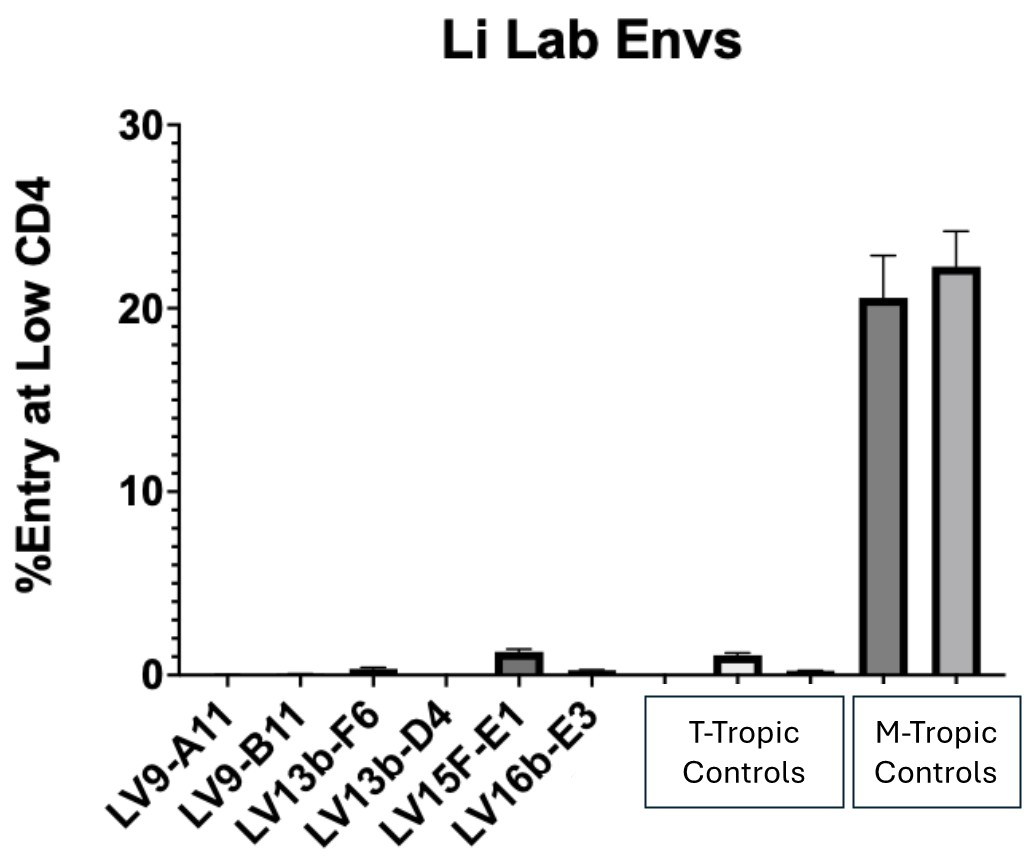

**Supplemental Figure 7.** Tropism phenotyping of two secondary and two primary NSV participants. LV13 and LV15 were primary NSV, while LV9 and LV16 were secondary NSV participants. The suffixes (e.g., A11, B11, F6) indicate individual *Env* clones derived from distinct wells.

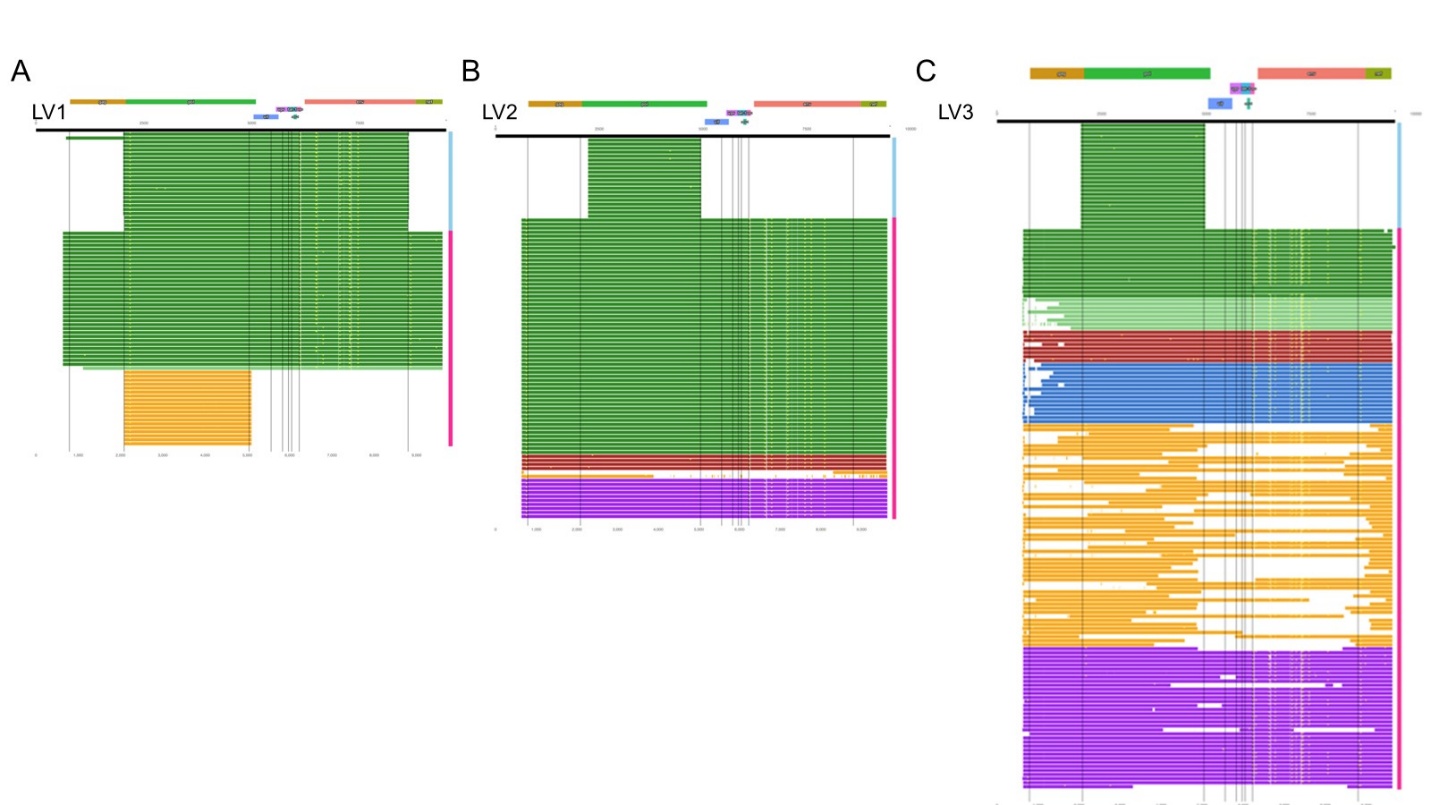

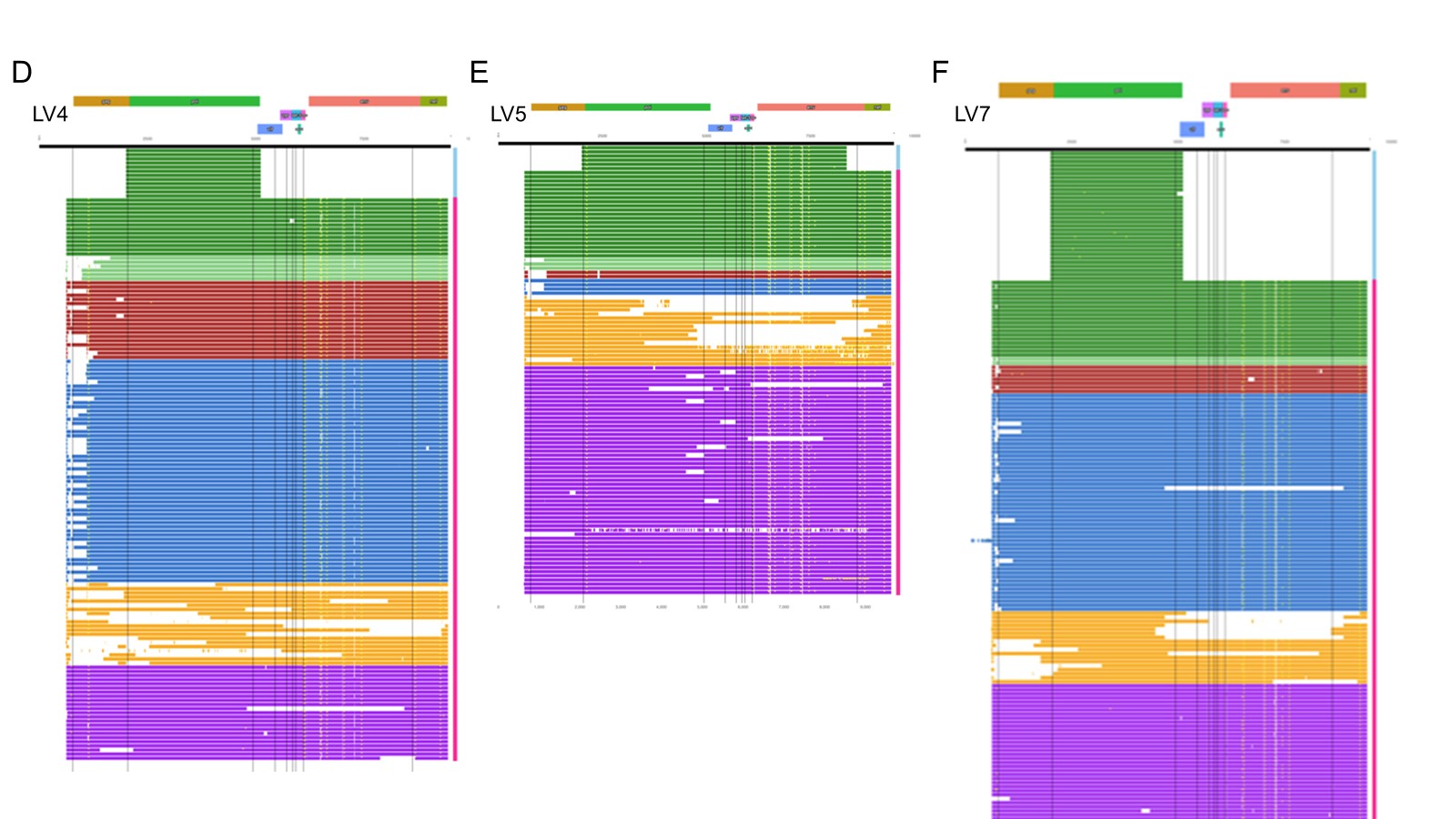

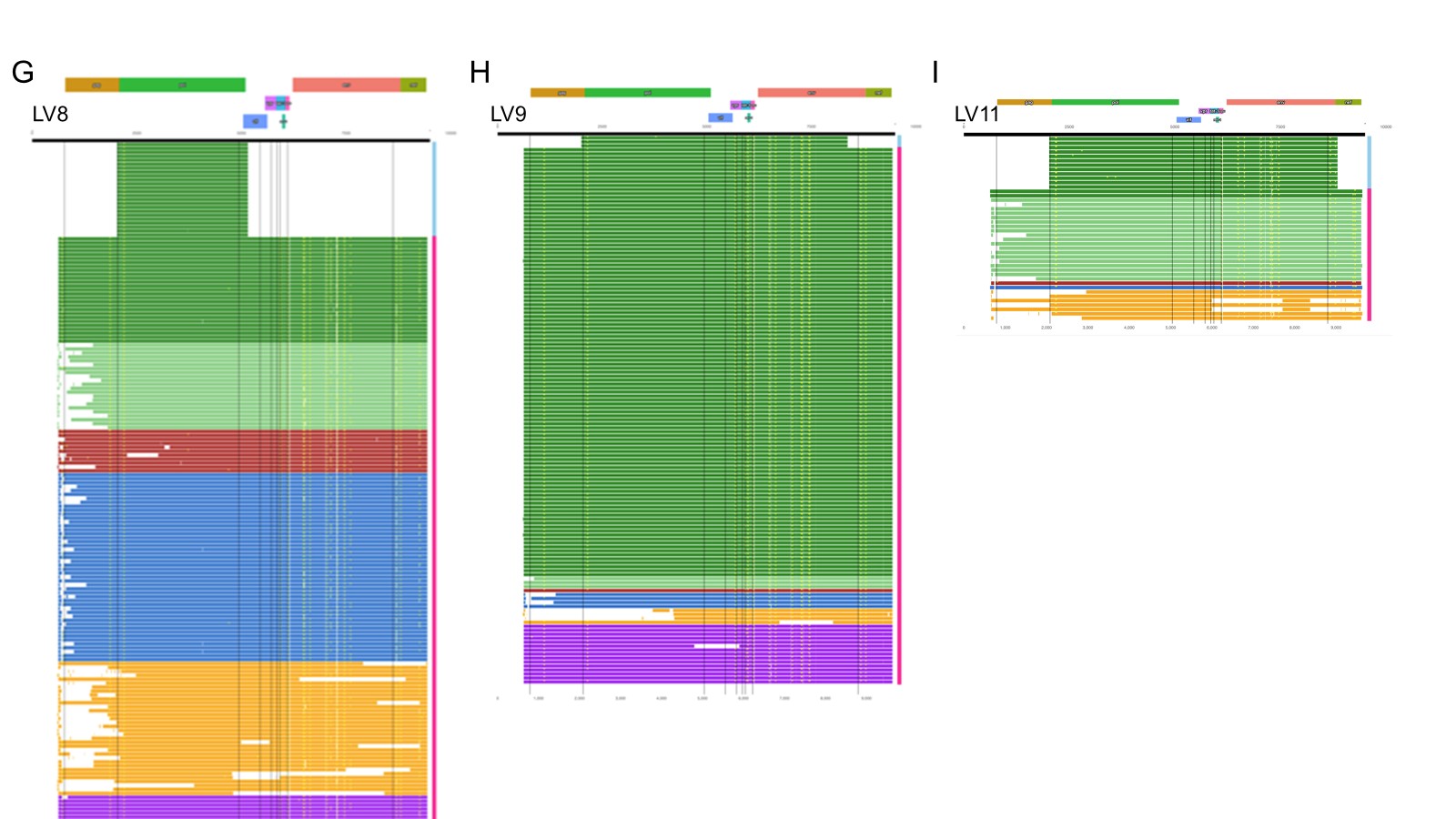

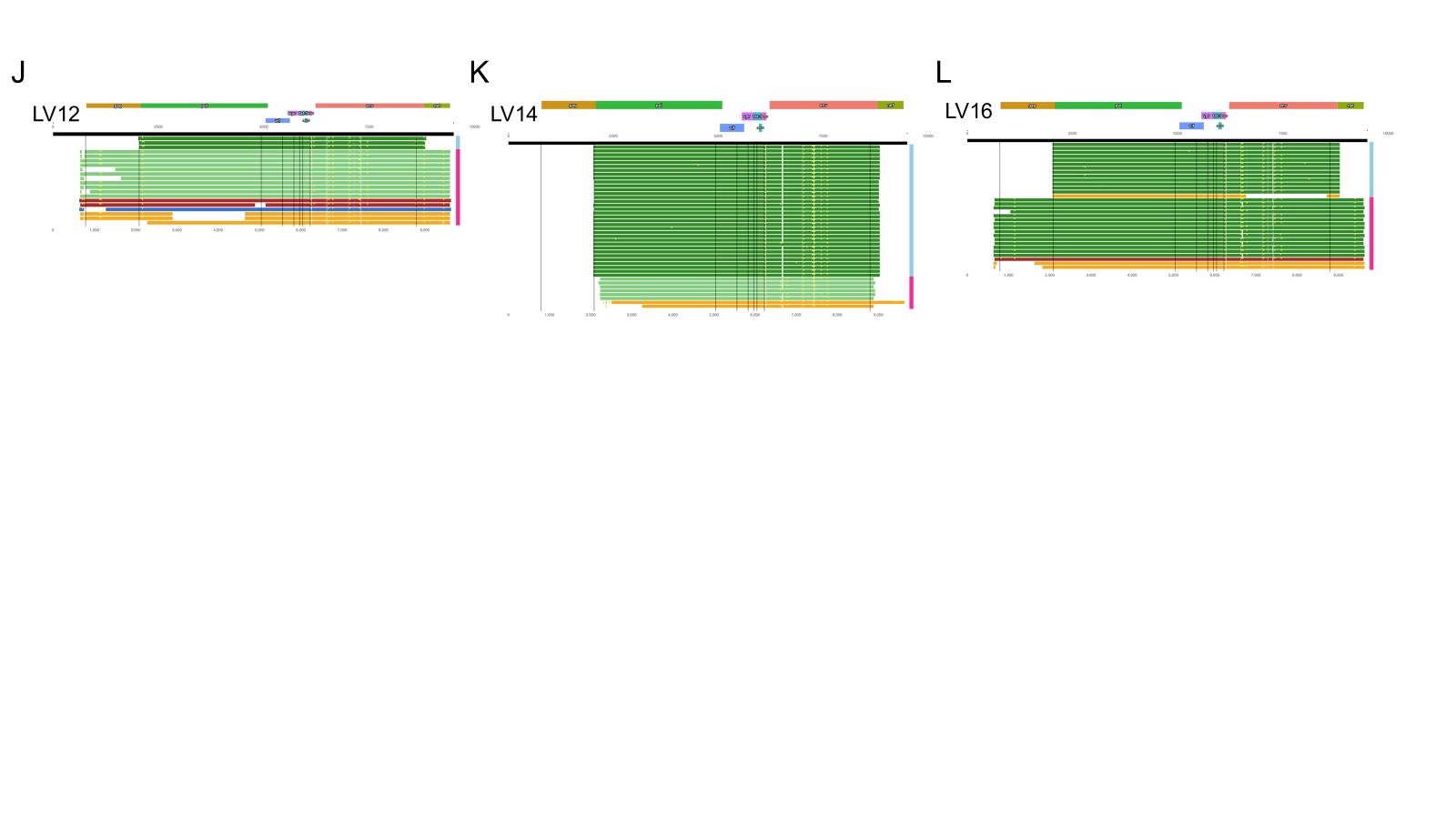

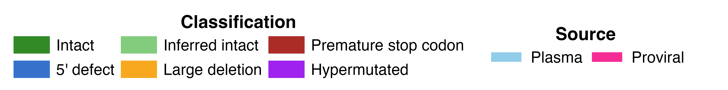

**Supplemental Figure 8.** Virogram plot of additional secondary NSV participants. Side bar shows sequence source plasma (blue) or proviral (pink). Color of bar indicates sequence intactness. Total length of bar shows sequenced region with plasma sequences spanning either *pol-env* or *pol*, and proviral sequences are near-full length.

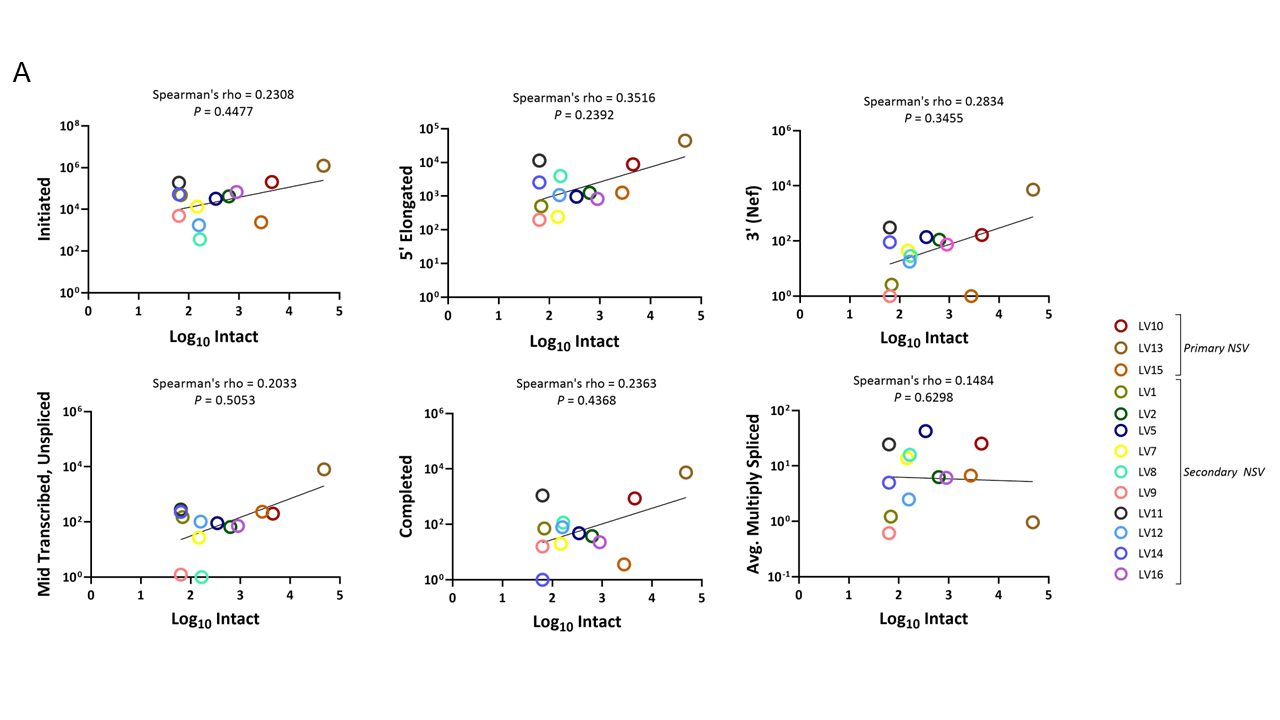

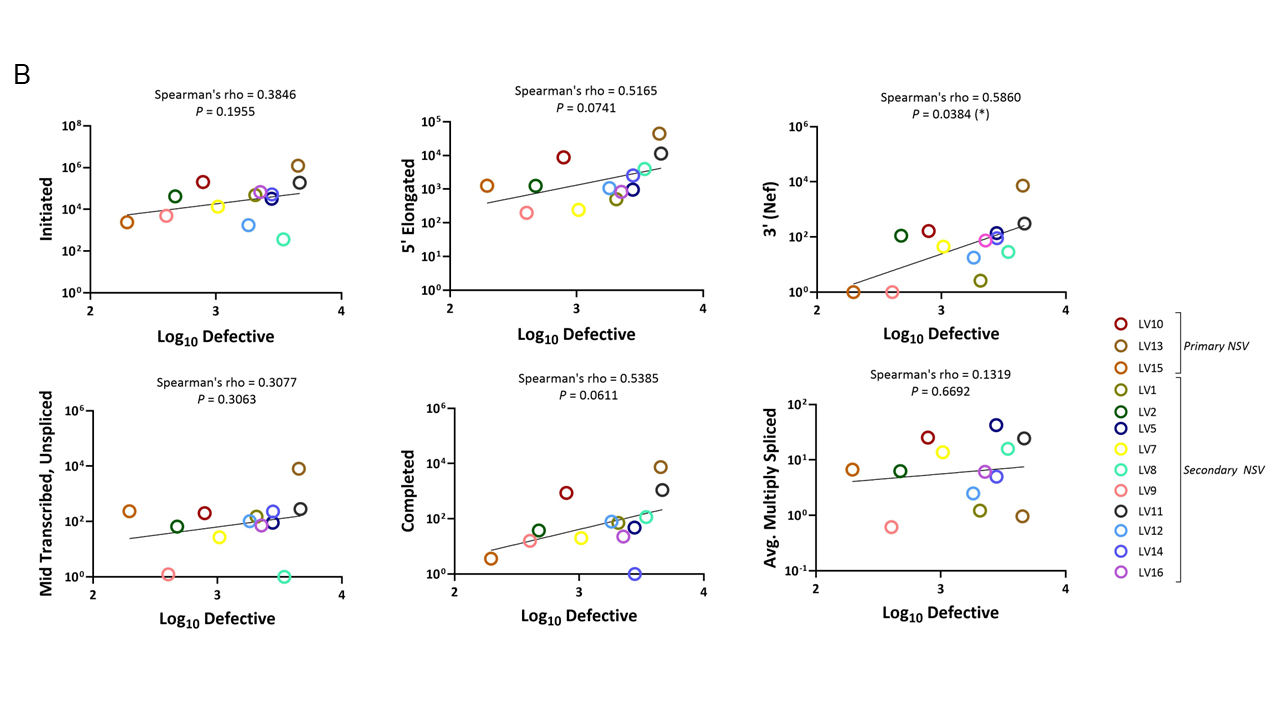

**Supplemental Figure S9.** Correlation between RNA transcription levels and (A) intact reservoir size and (B) defective reservoir size. Correlations were assessed using Spearman’s rank correlation coefficient (ρ).

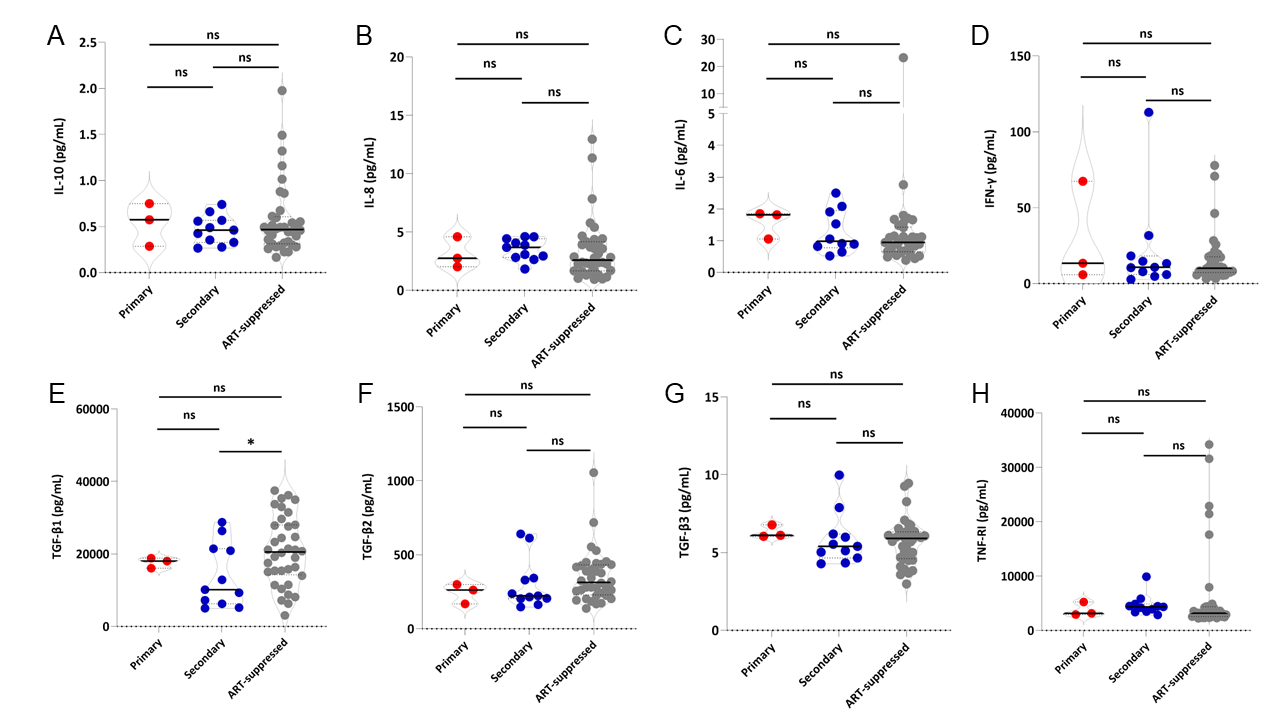

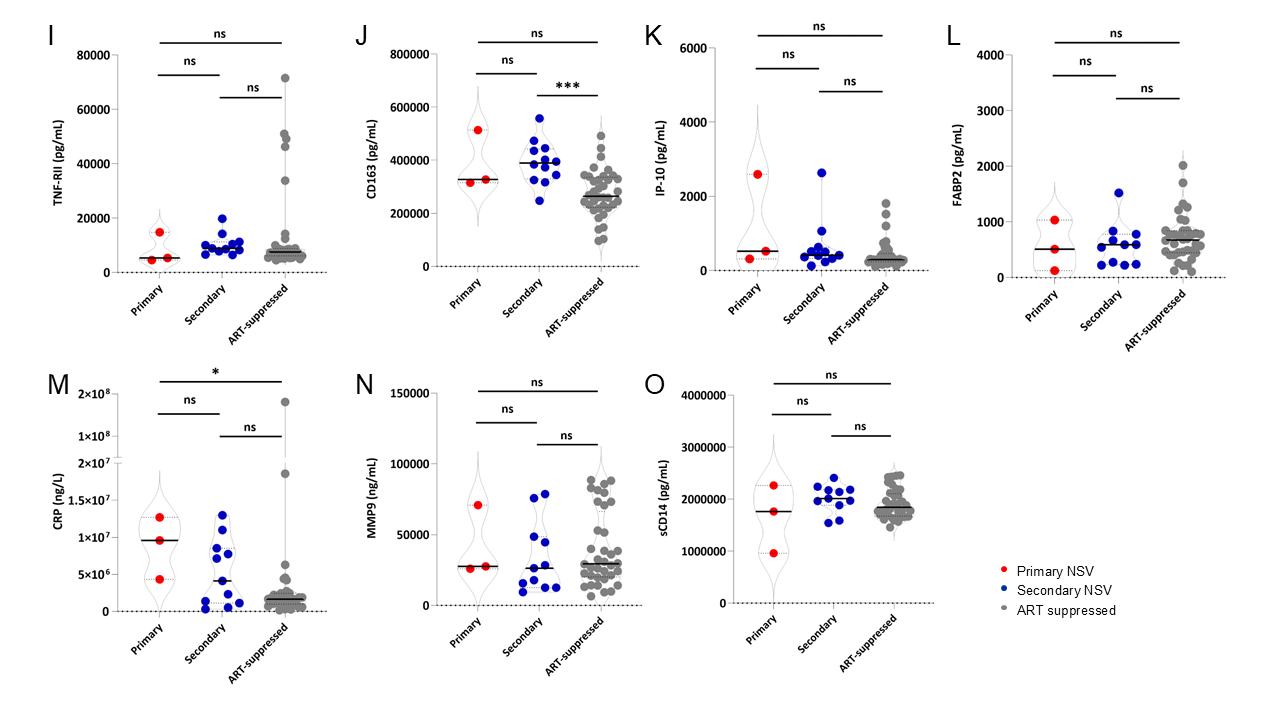

**Supplemental Figure S10.** Soluble inflammatory markers in primary versus secondary NSV participants. (A) IL-10, (B) IL-8, (C)) IL-6, (D) IFN-γ, (E) TGF-β1, (F) TGF-β2, (G) TGF-β3, (H) TNF-RI, (I) TNF-RII, (J) CD163, (K) IP-10, (L) FABP2, (M) CRP, (N) MMP9, and (O) soluble CD14 in primary NSV, secondary NSV, and ART-suppressed cohorts. Center lines indicated median levels and dotted lines indicated the first and third quartiles. Comparisons were performed by Mann–Whitney *U* test.

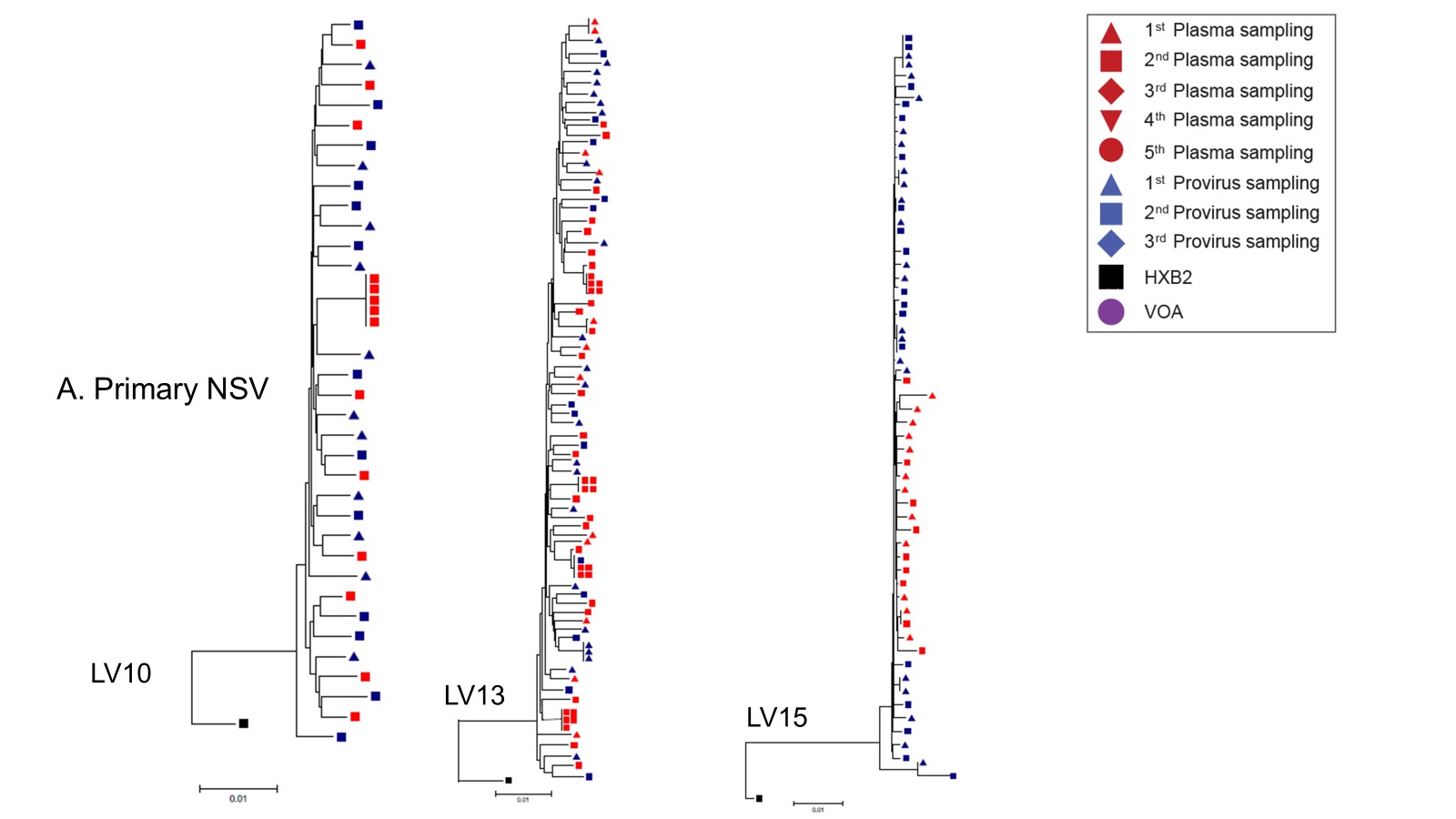

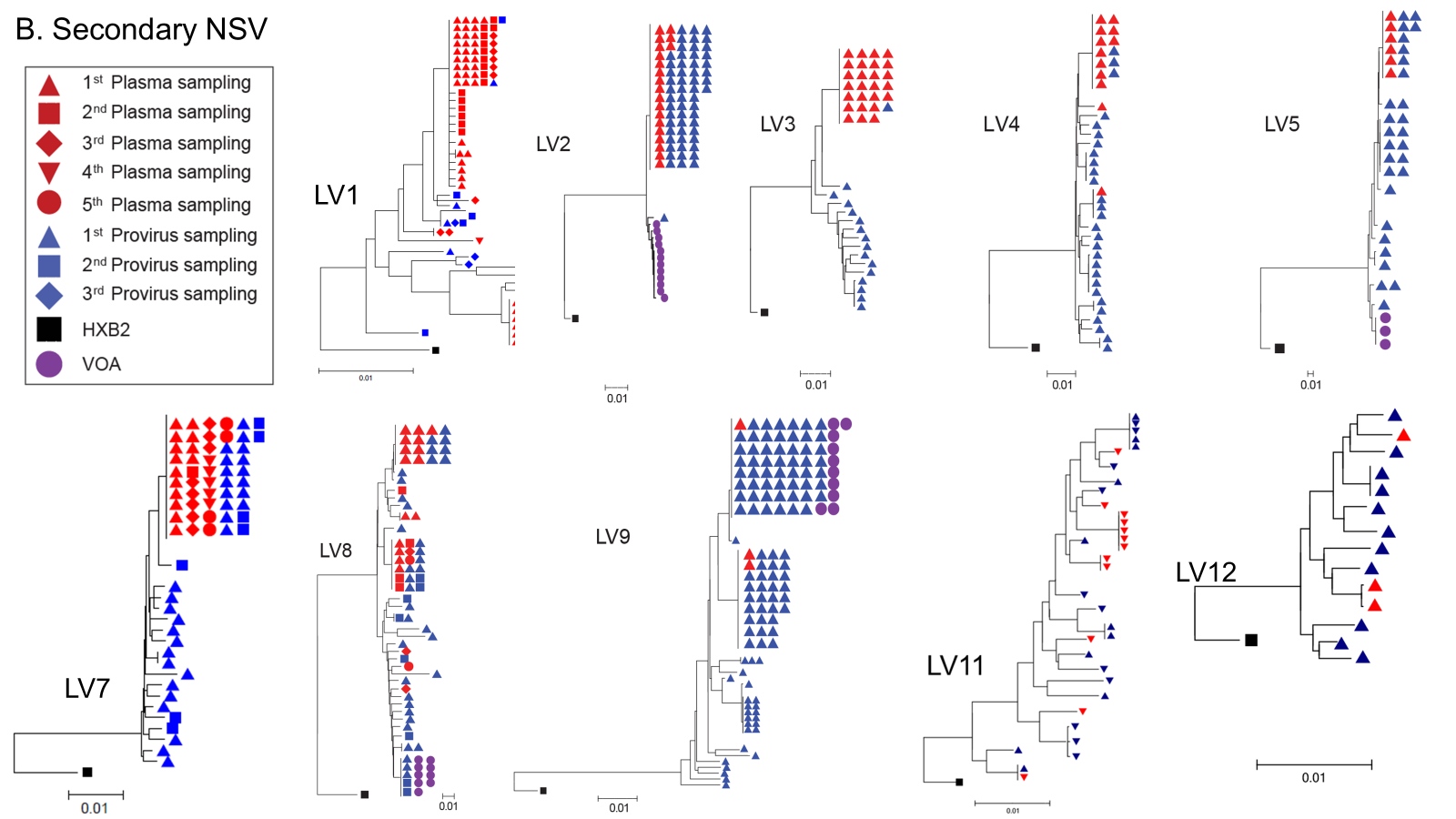

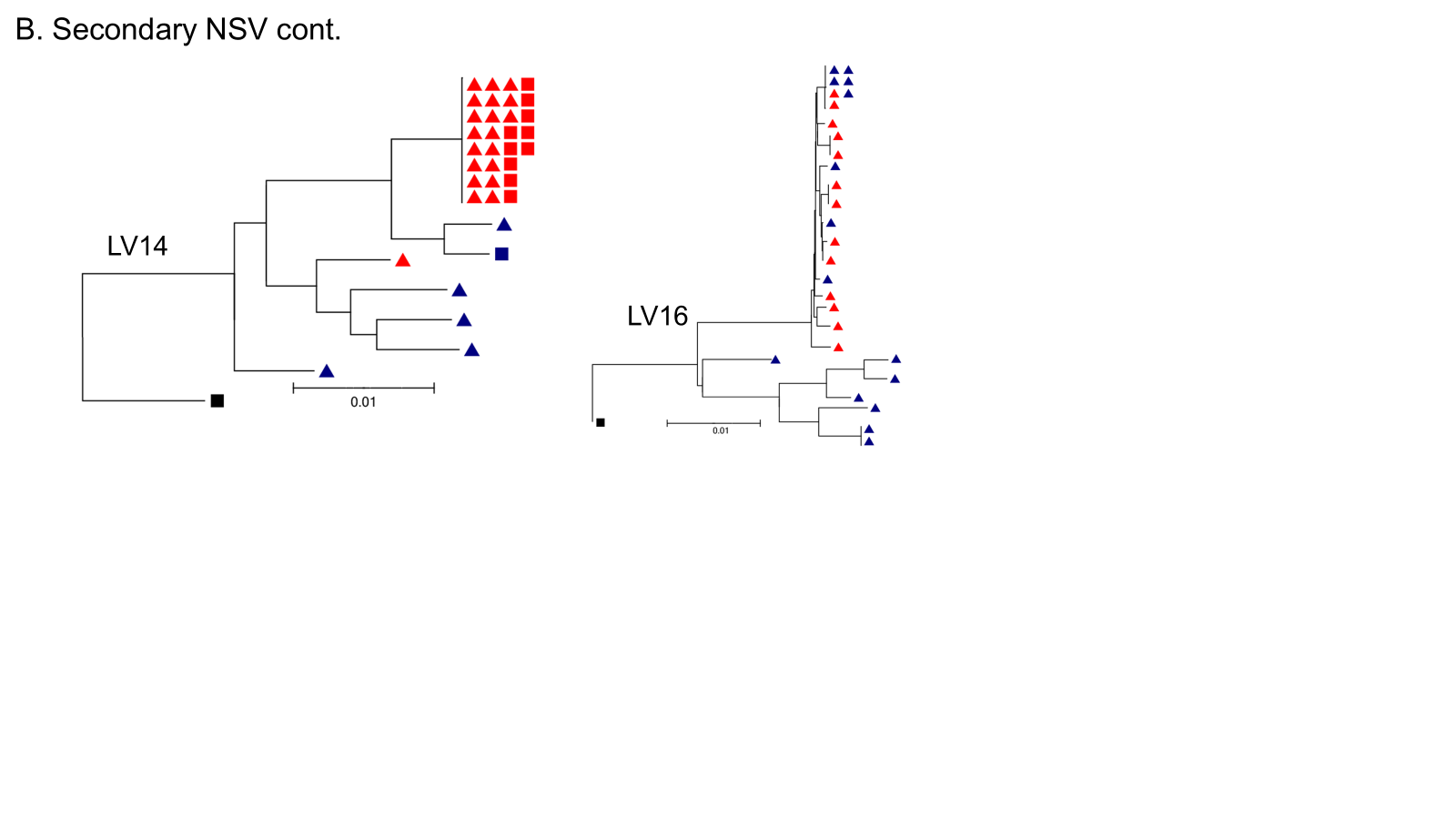

**Supplemental Figure S11.** Individual phylogenetic neighbor joining trees of primary and secondary NSV participants. Sequences are derived from provirus (blue), plasma (red), or viral outgrowth assay (purple). The timepoint of the sequence is indicated by the shape.

**Supplementary Table 1.** Clinical characteristics of non-suppressible viremia participants

| ID | Type of NSV | Age group^A^ | Sex | Race | CD4^+^ T-cell nadir  (cells/mm^3^) | CD4^+^ T-cell count,  the latest (cells /mm^3^) | ART regimen^A^ | GSS^A^ |
| --- | --- | --- | --- | --- | --- | --- | --- | --- |
| LV1 | Secondary | 60-70 | M | Hispanic | 149 | 462 | TDF/FTC/DTG/DRV/r | 2.25 |
| LV2 | Secondary | 50-60 | M | Native American | 360 | 680 | TAF/FTC/DTG/MVC/DRV/c | 5 |
| LV3 | Secondary | 60-70 | M | White | 531 | 531 | TAF/FTC/DTG | 2.25 |
| LV4 | Secondary | 50-60 | M | White | 820 | 1472 | ABC/3TC/DTG | 3 |
| LV5 | Secondary | 60-70 | M | African American | 276 | 598 | TAF/FTC/DOR/BIC/DRV/r | 2.25 |
| LV7 | Secondary | 60-70 | M | White | 68 | 647 | TAF/FTC/BIC | 3 |
| LV8 | Secondary | 40-50 | F | African American | 208 | 1423 | TAF/FTC/BIC | 2 |
| LV9 | Secondary | 50-60 | M | White | 577 | 927 | TAF/FTC/BIC | 2 |
| LV10 | Primary | 30-40 | M | White | 24 | 647 | TAF/FTC/DTG/DRV/r | 4 |
| LV11 | Secondary | 60-70 | M | White | 170 | 258 | TAF/FTC/BIC | 2 |
| LV12 | Secondary | 40-50 | M | African American | 112 | 876 | TAF/FTC/BIC | 2.5 |
| LV13 | Primary | 20-30 | F | White | 20 | 487 | TAF/FTC/BIC | 2.75 |
| LV14 | Secondary | 70-80 | M | White | 285 | 412 | TAF/FTC/RPV/DRV/c | 4 |
| LV15* | Primary | 20-30 | F | African American | 310 | 1517 | RPV/CAB | 2 |
| LV16 | Secondary | 50-60 | F | African American | 30 | 481 | TAF/FTC/RPV/DRV/c | 4 |
| Median | - | 55 | - | - | 208 | 647 | - | 2.5 |

^A^at the time of initial sample collection. *Perinatal infection.

NSV, non-suppressible viremia; ART, antiretroviral therapy; GSS, genotypic susceptibility score; TDF, tenofovir disoproxil fumarate; FTC, emtricitabine; DTG, dolutegravir; DRV/r, darunavir-ritonavir; TAF, tenofovir alafenamide; MVC,maraviroc; DRV/c, darunavir-cobicistat; ABC, abacavir; 3TC, lamivudine; DOR, doravirine; BIC, bictegravir; RAL, raltegravir; ATV, atazanavir; RPV, rilpivirine; CAB, cabotegravir

**Supplementary Table 2.** Antiretroviral drug concentrations measured by liquid chromatography for NSV participants

| ID | ART Regimen | Plasma concentration (ng/mL) | | | | | | | | | | | |
| --- | --- | --- | --- | --- | --- | --- | --- | --- | --- | --- | --- | --- | --- |
|  |  | TP1 | | | | | | TP2 | | | | | |
|  |  | DTG | DRV | CAB | | RPV | | DTG | | DRV | | CAB | RPV |
| LV1 | TDF/FTC/DTG/DRV/r | 290 | 1520 | | - | | - | | 2880 | | 7210 | - | - |
| LV2 | TAF/FTC/DTG/MVC/DRV/c | 1520 | 6270 | | - | | - | | - | | - | - | - |
| LV15 | RPV/CAB | - | - | | 1307 | | 37.5 | | - | | - | 1350 | 57 |

NSV, non-suppressible viremia; ART, antiretroviral therapy; TDF, tenofovir disoproxil fumarate; FTC, emtricitabine; DTG, dolutegravir; DRV/r, darunavir-ritonavir; TAF, tenofovir alafenamide; MVC,maraviroc; DRV/c, darunavir-cobicistat; RPV, rilpivirine; CAB, cabotegravir; TP, timepoint
